# A trans-ancestry genome-wide association study of unexplained chronic ALT elevation as a proxy for nonalcoholic fatty liver disease with histological and radiological validation

**DOI:** 10.1101/2020.12.26.20248491

**Authors:** Marijana Vujkovic, Shweta Ramdas, Kimberly M. Lorenz, Xiuqing Guo, Rebecca Darlay, Heather J. Cordell, Jing He, Yevgeniy Gindin, Chuhan Chung, Rob P Meyers, Carolin V. Schneider, Joseph Park, Kyung M. Lee, Marina Serper, Rotonya M. Carr, David E. Kaplan, Mary E. Haas, Matthew T. MacLean, Walter R. Witschey, Xiang Zhu, Catherine Tcheandjieu, Rachel L. Kember, Henry R. Kranzler, Anurag Verma, Ayush Giri, Derek M. Klarin, Yan V. Sun, Jie Huang, Jennifer Huffman, Kate Townsend Creasy, Nicholas J. Hand, Ching-Ti Liu, Michelle T. Long, Jie Yao, Matthew Budoff, Jingyi Tan, Xiaohui Li, Henry J. Lin, Yii-Der Ida Chen, Kent D. Taylor, Ruey-Kang Chang, Ronald M. Krauss, Silvia Vilarinho, Joseph Brancale, Jonas B. Nielsen, Adam E. Locke, Marcus B. Jones, Niek Verweij, Aris Baras, K. Rajender Reddy, Brent A. Neuschwander-Tetri, Jeffrey B. Schwimmer, Arun J. Sanyal, Naga Chalasani, Katherine A. Ryan, Braxton D. Mitchell, Dipender Gill, Andrew D. Wells, Elisabetta Manduchi, Yedidya Saiman, Nadim Mahmud, Donald R. Miller, Peter D. Reaven, Lawrence S. Phillips, Sumitra Muralidhar, Scott L. DuVall, Jennifer S. Lee, Themistocles L. Assimes, Saiju Pyarajan, Kelly Cho, Todd L. Edwards, Scott M. Damrauer, Peter W. Wilson, J. Michael Gaziano, Christopher J. O’Donnell, Amit V. Khera, Struan F.A. Grant, Christopher D. Brown, Philip S. Tsao, Danish Saleheen, Luca A. Lotta, Lisa Bastarache, Quentin M. Anstee, Ann K. Daly, James B. Meigs, Jerome I. Rotter, Julie A. Lynch, Regeneron Genetics Center, DiscovEHR Collaboration, EPoS Consortium Investigators, VA Million Veteran Program, Daniel J. Rader, Benjamin F. Voight, Kyong-Mi Chang

## Abstract

Nonalcoholic fatty liver disease (NAFLD) is a growing cause of chronic liver disease. Using a proxy NAFLD definition of chronic alanine aminotransferase elevation (cALT) without other liver diseases, we performed a trans-ancestry genome-wide association study in the Million Veteran Program including 90,408 cALT cases and 128,187 controls. In the Discovery stage, seventy-seven loci exceeded genome-wide significance – including 25 without prior NAFLD or ALT associations – with one additional locus identified in European-American-only and two in African-American-only analyses (P<5×10^-8^). External replication in cohorts with NAFLD defined by histology (7,397 cases, 56,785 controls) or liver fat extracted from radiologic imaging (n=44,289) validated 17 SNPs (P<6.5×10^-4^) of which 9 were novel (*TRIB1*, *PPARG*, *MTTP*, *SERPINA1*, *FTO*, *IL1RN*, *COBLL1*, *APOH*, and *IFI30*). Pleiotropy analysis showed that 61 of 77 trans-ancestry and all 17 validated SNPs were jointly associated with metabolic and/or inflammatory traits, revealing a complex model of genetic architecture. Our approach integrating cALT, histology and imaging reveals new insights into genetic liability to NAFLD.

## Introduction

Chronic liver disease with progression to cirrhosis and hepatocellular carcinoma is a global health issue^1^. In particular, nonalcoholic fatty liver disease (NAFLD) – a hepatic phenotype associated with metabolic syndrome and insulin resistance – is an increasingly common cause of chronic liver disease with an estimated world prevalence of 25% among adults^1–5^. In the United States (US), NAFLD prevalence is projected to reach 33.5% among the adult population by 2030, due in large part to rising rates of obesity and other cardiometabolic risk factors^6^. NAFLD is defined by ≥5% fat accumulation in the liver (hepatic steatosis) in the absence of other known causes for liver disease, based on liver biopsy and/or non-invasive radiologic imaging^3, 4^. The clinical spectrum of NAFLD ranges from bland steatosis to nonalcoholic steatohepatitis (NASH) involving inflammation and hepatocellular ballooning injury with progressive fibrosis. At least 20% of patients with NAFLD develop NASH with increased risk of consequent cirrhosis and primary liver cancer^5, 6^. To date, there is no licensed drug approved to treat NAFLD and prevent its progression, and the general therapeutic approach focuses on improving the underlying metabolic disorders such as glucose control and promotion of weight loss.

Individual susceptibility to NAFLD involves both genetic and environmental risk factors. Current estimates of NAFLD heritability range from 20% to 50%^7^ while risk factors for NAFLD include obesity (in particular, abdominal adiposity), insulin resistance and several features of metabolic syndrome^2, 5, 6, 8^. Several genetic variants that promote the full spectrum of fatty liver disease have been identified in genome-wide association studies (GWAS) utilizing cohorts based on liver biopsy, imaging, and/or isolated liver enzyme values^9–22^. The most prominent variants include p.I148M in *PNPLA3* and p.E167K in *TM6SF2*, which increase NAFLD risk, and a loss-of-function variant in *HSD17B13* that confers protection against NASH^16^. However, the limited number of genetic associations in NAFLD contrasts with other cardiometabolic disorders where hundreds of loci have been mapped to date, such as obesity^23, 24^, type 2 diabetes^25^, hypertension^26^, and plasma lipids^27^. This highlights the need for expanded discovery with larger samples and greater population diversity, with further integration of functional genomics data sets to potentially identify the effector genes^28^.

The Million Veteran Program (MVP) is among the world’s largest and most ancestrally diverse biobanks^29^. The availability of comprehensive, longitudinally collected Veterans Health Administration (VA) electronic health records for US Veteran participants in the MVP also makes it a promising resource for precision medicine. However, NAFLD is markedly underdiagnosed clinically due to the invasive nature of the liver biopsy procedure, variable use of imaging modalities and poor sensitivity of international diagnostic codes (ICD) for NAFLD^4, 30^. The use of chronic elevation of serum alanine aminotransferase levels (cALT) as a proxy for NAFLD was shown to improve specificity and positive predictive value in the NAFLD diagnostic algorithm within the VA Corporate Data Warehouse^31^. Accordingly, we recently adapted and validated a cALT phenotype as a proxy for NAFLD to facilitate case identification in MVP^21^, applying a rigorous exclusion of other conditions that are known to increase liver enzymes (e.g., viral hepatitis, alcoholic liver disease, autoimmune liver disease, and known hereditary liver disease). Moreover, we showed that our cALT phenotype had a sensitivity of 80%, a specificity of 89%, an accuracy of 85%, a positive predictive value of 89%, and an area-under-the-curve of 87% compared to gold standard abdominal imaging, liver biopsy, and clinical notes in a well-characterized sample of 178 patients in the VA healthcare system. In the current study, we applied this cALT phenotype in MVP participants of 4 ancestral groups^32^, and identified 90,408 NAFLD cases and 128,187 controls (**Figure 1,** and **Supplementary Figure 1**).

**Figure 1.**
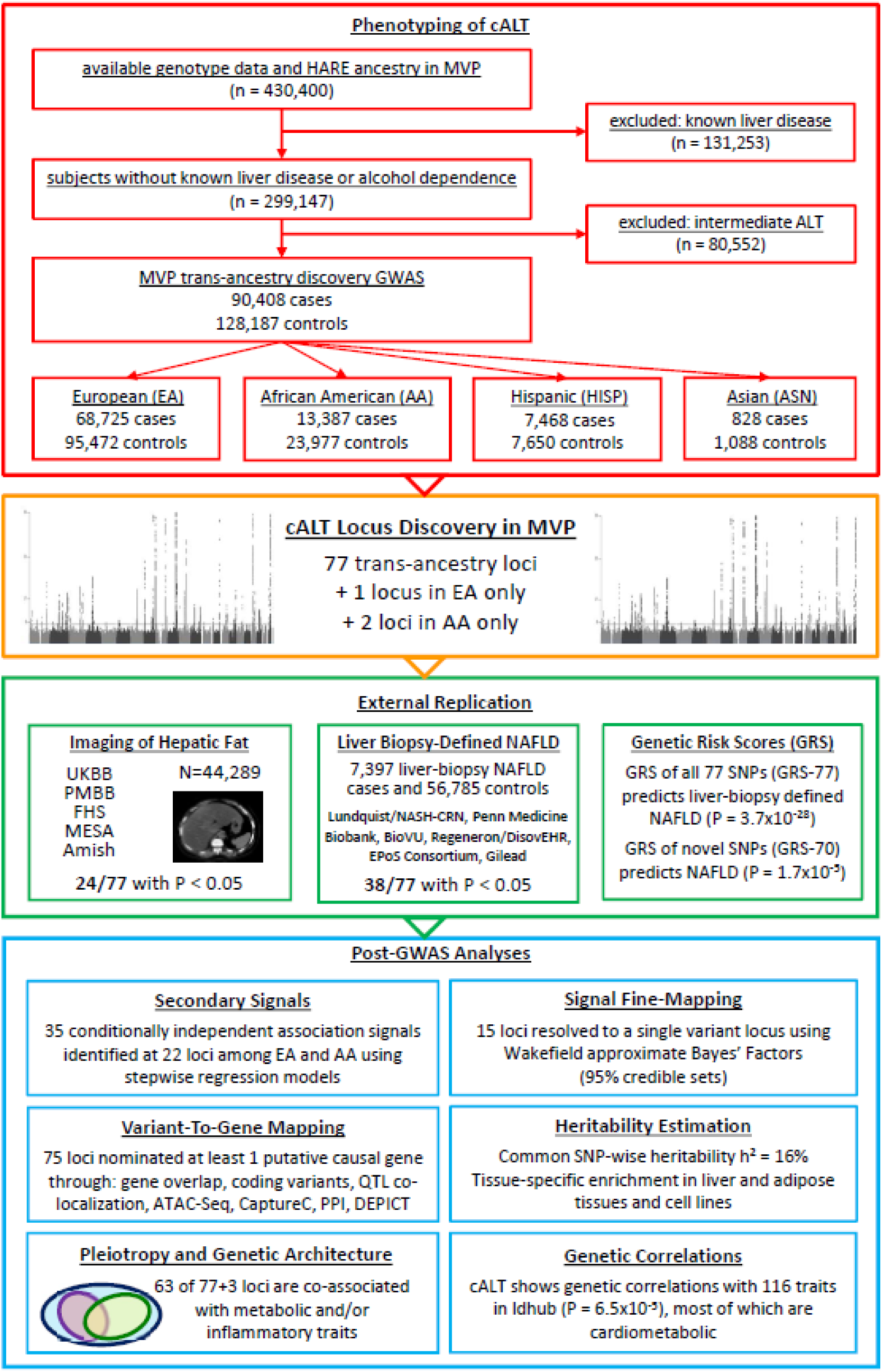
Overview of analysis pipeline. The flow diagram shows in the red box our study design with initial inclusion of 430,400 Million Veteran Program participants with genotyping and ancestry classification by HARE, exclusion of individuals with known liver disease or alcohol dependence and inclusion of subjects based on chronic ALT elevation (case) or normal ALT (control). This resulted in 90,408 NAFLD cases and 128,187 controls with EA, AA, HISP and ASN ancestries that were examined in primary trans-ancestry and ancestry-specific genome-wide association scans. The orange box of the flow diagram highlights our results of trans-ancestry and ancestry-specific meta-analyses identifying 77 trans-ancestry loci + 1 EA-specific + 2 AA-specific loci that met genome-wide significance. The green box summarizes the results from external replication cohorts, whereas the blue box indicates all the post-GWAS annotation analyses that we performed, which include secondary signal analysis, fine-mapping (95% credible sets), (tissue-specific) heritability estimation, genetic correlations analysis, variant-to-gene mapping and pleiotropy analysis.

Our aims are to: (i) perform a trans-ancestry genetic susceptibility analysis of the cALT phenotype in MVP; (ii) replicate the lead SNPs in external NAFLD cohorts with hepatic fat defined by liver histology or radiologic imaging; (iii) identify putative causal genes at the lead loci by using an ensemble “variant-to-gene” mapping method that integrates results from coding and functional genomic annotations, and quatitative trait loci (QTL); and (iv) describe the genetic architecture at each associated locus by characterizing the profile of additional phenotypic associations using data from the GWAS catalog, MVP LabWAS, and MRC IEU OpenGWAS database.

## Results

### An ancestry-diverse study population with a high prevalence of cardiometabolic traits

Our study consisted of 90,408 cALT cases and 128,187 controls comprising four ancestral groups, namely European-Americans (EA, 75.1%), African-Americans (AA, 17.1%), Hispanic-Americans (HISP, 6.9%), and Asian-Americans (ASN, 0.9%, **Supplemental Table 1**, **Supplemental Figure 1**). Consistent with the US Veteran population, MVP cases and controls (n = 218,595) were predominantly male (92.3%) with an average age of 64 years at study enrollment (**Supplemental Table 1**). The prevalence of cirrhosis and advanced fibrosis in the cALT cases ranged from 4.7% to 9.1%. With the exclusion of other known causes of liver disease as described previously^21^, cases were enriched for metabolic disorders (type 2 diabetes, hypertension, and dyslipidemia), suggesting a link between chronic ALT elevation and metabolic risk factors.

### Identification of trans-ancestry and ancestry-specific cALT-associated loci in MVP

To identify genetic loci associated with cALT, we performed a trans-ancestry genome-wide scan by meta-analyzing summary statistics derived from each individual ancestry (**Methods** and **Figure 1**). In this trans-ancestry scan, 77 independent sentinel SNPs met the conventional genome-wide significance level (P < 5×10^-8^), of which 60 SNPs exceeded trans-ancestry genome-wide significance (P < 5×10^-9^). Of the 77 SNPs, 52 were previously reported to be associated with ALT, including 9 that were also associated with NAFLD (i.e., *PNPLA3*, *TM6SF2*, *HSD17B13*, *PPP1R3B*, *MTARC1*, *ERLIN1*, *APOE*, *GPAM,* and *SLC30A10;LYPLAL1*; **Figure 2 and Supplemental Table 2**)^9-12, 14, 16–22, 33–35^. Of the 25 newly identified loci, 14 were novel (*IL1RN*, *P2RX7*, *CASP8*, *MERTK*, *TRPS1*, *OGFRL1*, *HLA*, *SMARCD2;DDX42*, *CRIM1*, *FLT1*, *DNAJC22*, *HKDC1*, *UHRF2*, *STAP2;MPND*), whereas 11 have been associated with gamma-glutamyl transferase (GGT) and/or alkaline phosphatase (ALP) levels^35^.

**Figure 2.**
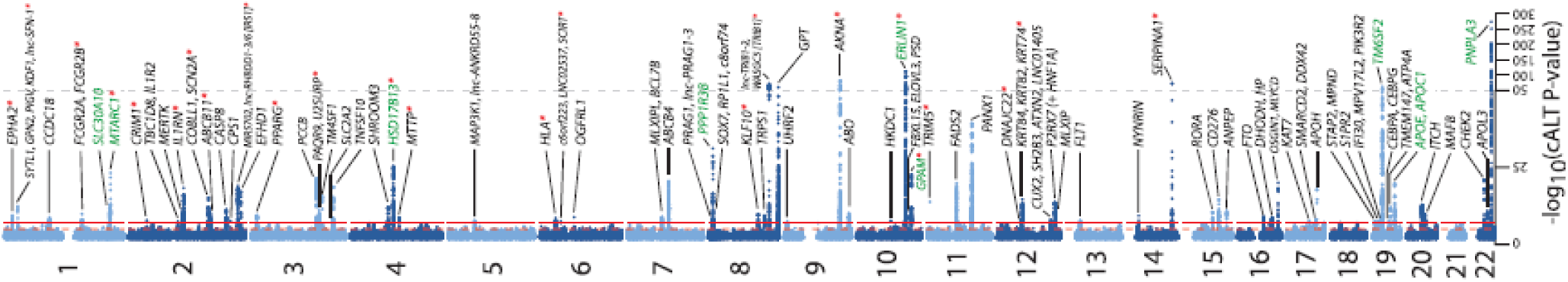
Manhattan plot of NAFLD GWAS of 90,408 NAFLD and 128, 187 controls in trans-ancestry meta-analysis. Nominated genes are indicated for 77 loci reaching genome-wide significance (P<5×10^-8^). Previously reported NAFLD-loci with genome-wide significant association are indicated in green font. The red stars indicate the SNPs that have been validated with liver biopsy and/or radiologic imaging.

In the ancestry-specific analyses, 55 loci in EAs, eight loci in AAs, and three in HISPs exceeded conventional genome-wide significance (P < 5×10^-8^, **Supplemental Tables 3-5** and **Supplemental Figures 2-4**), of which 1 EA SNP and 2 AA SNPs were not captured in the trans-ancestry analysis. No variants among ASN subjects achieved genome-wide significance, likely due to limited sample size (**Supplemental Figure 5**). Notably, the top two SNPs in the AA-only scan (with the genes *GPT* and *ABCB4* nearby each SNP, respectively) showed stronger associations than observed for *PNPLA3.* These two SNPs are polymorphic among AA’s but nearly monomorphic in all other populations.

### Replication of cALT-associated loci in liver biopsy and radiologic imaging data

To validate that our cALT-associated SNPs capture genetic susceptibility to NAFLD, we assembled two external NAFLD cohorts, namely: (i) a **<u>Liver Biopsy Cohort</u>** consisting of 7,397 histologically characterized NAFLD cases and 56,785 population controls from various clinical studies **(Supplemental Table 6-7**), and (ii) a **<u>Liver Imaging Cohort</u>** consisting of 44,289 participants with available radiologic liver imaging data and quantitative hepatic fat (qHF) measurements (**Supplemental Table 8**). For each cohort, we performed a trans-ancestry lookup of the 77 lead SNPs (**Methods**). In the Liver Biopsy Cohort, there was directional concordance between effect estimates of biopsy-defined NAFLD in 66 of 77 SNPs (86%), including 15 SNPs with a significant association (adjusted Bonferroni nominal P < 6.5×10^-4^), of which 8 have not been reported in genome- or exome-wide association studies previously (e.g., *TRIB1*, *MTTP*, *APOH*, *IFI30*, *COBLL1*, *SERPINA1*, *IL1RN*, and *FTO*, **Supplemental Table 7**)^12^. In the Liver Imaging Cohort, there was directional concordance between effect estimates of qHF in 49 of 77 SNPs (64%). Among these, 11 were significantly associated with qHF (adjusted Bonferroni nominal P < 6.5×10^-4^**, Supplemental Table 8**) of which 6 were novel (e.g., *TRIB1*, *MTTP*, *APOH*, *IFI30*, *COBLL1*, and *PPARG*). As observed in previous studies owing to its role in glycogen storage (and associated impact on imaging)^12, 36^, the *PPP1R3B* locus was significantly associated with qHF, however in the opposite direction from cALT, and not associated with biopsy-proven NAFLD. Collectively, 17 of 77 SNPs were validated in external histologic and/or radiologic NAFLD, of which nine were previously unreported; namely, five in both biopsy and imaging cohorts (*TRIB1, MTTP, APOH, IFI30*, and *COBLL1*), three in biopsy cohort alone (*FTO, SERPINA1*, and *IL1RN*) and one in imaging cohort with a nominal significance in biopsy cohort (*PPARG*) (**Supplemental Table 2**). An additional 24 SNPs were nominally associated (P < 0.05) with directional concordance with histological and/or radiologic hepatic fat.

We performed a SNP-specific statistical power analysis to investigate the SNP-specific type I error (α) at a P-value of = 6.5×10^-4^ (Bonferroni) using the respective effect estimates and allele frequencies (**Supplemental Table 2 and 7**). A total of 22 loci showed sufficient statistical power (> 80%) for replication in the liver biopsy cohort, of which 10 replicated **(Supplemental Figure 6)**. Despite sufficient power, 12 SNPs did not replicate in the liver biopsy cohort, which included *GPT*, *PPP1R3B*, *OGFRL1*, *TNFSF10*, and *PANX1*. Remarkably, in the group of 55 SNPs without sufficient statistical power for replication, 6 SNPs did in fact replicate (namely PPARG, MTTP, FTO, IL1RN, IFI30, and COBLL1). We note that for cALT SNPs which were replicated in the histological NAFLD cohort, their effect size in histological NAFLD cohort were, on average 92.4% higher than cALT effect estimate **(Supplemental Figure 7 and 8)**. Extrapolating to the remaining 49 SNPs without 80% statistical power for replication, 22 SNPs showed higher effect sizes for histological NAFLD than cALT and were therefore labeled as candidate NAFLD loci **(Supplemental Table 2)**.

### Genetic risk scores and histologically characterized NAFLD

We next constructed genetic risk scores (GRSs) based on effect estimates from our cALT GWAS SNPs in four independent liver biopsy cohorts to quantify the cumulative predictive power of our 77 sentinel variants (**Supplemental Table 9A, Supplemental Figure 9**). A 77 SNP-based GRS was predictive of NAFLD in the meta-analyzed liver biopsy cohorts (GRS-77, P = 3.7×10^-28^). Stratification of GRS-77 into a GRS consisting of a set of 9 well-established NAFLD SNPs (GRS-9), and a GRS consisting of 68 remaining SNPs (GRS-68) revealed significant independent capacity to predict histologically characterized NAFLD (GRS-9, P = 2.2×10^-11^; GRS-68, P = 2.4×10^-7^), further supporting the clinical relevance of our panel of SNPs derived from proxy NAFLD phenotype.

We further examined the SNPs in three groups based on their strength of external replication: (i) replicated at Bonferroni threshold; (ii) achieving nominal significance (P<0.05) with directional concordance without reaching Bonferroni significance; and (iii) P>0.05. As shown in **Supplemental Figure 10** and **Supplementary Table 9C**, a significant capacity for NAFLD prediction was noted not only for externally replicated SNPs (Beta: 0.644, P=2.53 x 10^-10^) but also the nominal SNPS set albeit with attenuated effect (Beta: 0.198, P=1 x 10^-4^).

### cALT heritability and genetic correlations with other phenotypes

To further characterize the architecture of our cALT phenotype and its relationship with other traits, we estimated heritability and genetic correlations with other traits using linkage disequilirium (LD) score regression^37–39^ (**Methods**). The SNP-based liability-scaled heritability was estimated at 16% (95% CI: 12-19, P < 1×10^-6^) in EA. Genetic correlation analysis between NAFLD from the EA-only scan and 774 complex traits from LD Hub **(Methods)** identified a total of 116 significant associations (adjusted Bonferroni nominal P < 6.5×10^-5^, **Supplemental Table 10**). These encompassed 78 cardiometabolic risk factors (67.2%) including measures of obesity and adiposity, type 2 diabetes, hypertension, dyslipidemia, and coronary artery disease, which is consistent with reports from observational studies correlating these traits to NAFLD^40^. Additional genetically correlated traits represented general health conditions (11.2%), educational attainment and/or socio-economic status (12.0%) and other conditions such as gastro-oesophageal reflux, osteoarthritis, gout, alcohol intake, smoking, ovary removal, and urinary albumin-to-creatinine ratio (9.5%).

### Identification of conditionally independent cALT-associated variants

To discover additional conditionally independent cALT signals within the 77 genomic regions, we performed exact conditional analysis using stepwise regression on individual level data for all single-ancestry sentinel variants (namely 51 EA, 8 AA, and 3 HISP genomic regions). We detected a total of 29 conditionally independent SNPs (P < 1×10^-5^) flanking five known and 16 novel cALT loci in EA (**Supplemental Table 11**). In particular, the *GPT* locus showed the highest degree of regional complexity with 4 conditionally independent SNPs, followed by *AKNA* with 3 conditionally independent SNPs. For one novel locus located on chromosome 12 between 121-122Mb, the trans-ancestry lead variant (rs1626329) was located in *P2RX7*, whereas the lead peak variant for EA mapped to *HNF1A* (rs1169292, **Supplemental Figure 11**). Both variants are in strong LD with distinct coding variants (*P2RX7*: rs1718119, Ala348Thr; *HNF1A*: rs1169288, Ile27Leu) and are compelling candidate genes for metabolic liver disease. In AA, we observed a total of six conditionally independent variants at three genomic regions, namely three variants at *GPT*, two at *AKNA* and one at the *ABCB4* locus (**Supplemental Table 11**). No conditionally independent variants were identified in Hispanic Americans. Collectively, 35 additional variants were identified at 22 loci across multiple ancestries by formal conditional analysis.

### Fine mapping to define potential causal variants in 95% credible sets

To leverage the increased sample size and population diversity to improve fine-mapping resolution, we computed statistically derived 95% credible sets using Wakefield’s approximate Bayes’ factors^41^ using the trans-ancestry, EA, AA, and HISP summary statistics (**Supplemental Table 12-15, Methods.** Trans-ancestry fine-mapping reduced the median 95% credible set size from 9 in EA (with IQR 3 - 17) to 7.5 variants (IQR 2 - 13). A total of 11 distinct cALT associations (e.g. *MTARC1, IL1RN, OGFRL1, PPP1R3B, SOX7;RP1L1;C8orf74, TRIB1, ERLIN1, TRIM5, OSGIN1;MLYCD, TM6SF2* and *PNPLA3*) were resolved to a single SNP in the trans-ancestry meta-regression, with an 4 additional loci suggesting single SNP sets from EA (n=2) and AA (n=2) ancestry-specific scans.

### Liver-specific enrichment of cALT heritability

To ascertain the tissues contributing to the disease-association underlying cALT heritability, we performed tissue-specific heritability analysis using stratified LD score regression. The strongest associations were observed for genomic annotations surveyed in liver, hepatocytes, adipose, and immune cell types among others (e.g., liver histone H3K36me3 and H3K4me1, adipose nuclei H3K27ac, spleen TCRγδ, eosinophils in visceral fat; P < 0.003, **Supplemental Table 16**). Medical subject heading (MeSH)-based analysis showed enrichment mainly in hepatocytes and liver (False Discovery Rate (FDR) < 5%, **Supplemental Table 17**). Gene set analysis showed strongest associations for liver and lipid-related traits (P-value < 1×10^-6^**, Supplemental Table 18**). Enrichment analyses using publicly-available epigenomic data implemented in GREGOR enrichment analysis (**Methods**) showed that most significant enrichments were observed for active enhancer chromatin state in liver, epigenetic modification of histone H3 in hepatocytes or liver-derived HepG2 cells (e.g. H3K27Ac, H3K9ac, H3K4me1, H3K4me3; adjusted Bonferroni nominal P < 1.8×10^-5^, **Supplemental Table 19 and 20**). These analyses additionally support the hypothesis that our cALT GWAS captures multiple physiological mechanisms that contribute to NAFLD heritability. Furthermore, DEPICT-based predicted gene function nominated 28 gene candidates, including the known genes *PNPLA3* and *ERLIN1* (FDR <5%, **Supplemental Table 21**), as well as well-known cardiometabolic disease genes such as *PPARG*.

### Coding variants in putative causal genes driving cALT associations

There were six novel trans-ancestry loci for which the lead SNP is a coding missense variant (**Supplementary Table 22**), namely Thr1412Asn in *CPS1*, Glu430Gln in *GPT*, Val112Phe in *TRIM5*, Ala163Thr in *DNAJC22*, Glu366Lys in *SERPINA1* and Cys325Gly in *APOH*. To identify additional coding variants that may drive the association between the lead SNPs and cALT risk, we investigated predicted loss of function (pLoF) and missense variants in high LD to the identified cALT lead variants (r^2^ > 0.7 in each respective 1000 Genomes super-population, **Supplemental Table 22**). Four previously described missense variants were replicated in the current study, including Thr165Ala in *MTARC1*, Ile291Val in *ERLIN1*, Glu167Lys in *TM6SF2* and Ile148Met in *PNPLA3*. Among novel loci, missense variants in high LD with lead variants included the genes *CCDC18, MERTK, APOL3, PPARG, MTTP, MLXIPL, ABCB4, GPAM, SH2B3, P2RX7, ANPEP, IFI30* and *MPV17L2*. Two additional missense variants were observed in *AKNA* and *NYNRIN*, however the coding variants were outside of the calculated boundaries of their respective credible sets. Among the trans-ancestry coding missense variants, eleven were predicted based on two methods (SIFT, PolyPhen-2) to have potentially deleterious and/or damaging effects in protein function (**Supplementary Table 22**)^42, 43^. An AA-specific locus on chromosome 7 (rs115038698, chr7:87024718) was in high LD to a nearby missense variant Ala934Thr in *ABCB4* (rs61730509, AFR r^2^=0.92) with a predicted deleterious effect, where the T-allele confers an increased risk of cALT (β=0.617, P=1.8×10^-20^). In summary, among our 77 trans-ancestry loci, 24 prioritized a candidate gene based on a missense variant in high LD with the lead SNP, including 5 novel loci with external validation (e.g., *SERPINA1*, *APOH*, *PPARG*, *MTTP* and *IFI30*;*MPV17L2*).

### Additional approaches to nominating putative causal genes

#### Co-localization analyses

We performed colocalization analyses with gene expression and splicing across 48 tissues measured by the GTEx project, and overlapped our lead SNPs with histone quantitative trait locus (QTL) data from primary liver to identify cALT-associated variants that are also associated with change in gene expression (eQTLs), splice isoforms (sQTLs), or histone modifications (hQTLs, **Methods**, **Supplemental Table 23**). Across all tissues, a total of 123 genes were prioritized, including 20 genes expressed in liver tissue (**Methods**). For liver tissue alone, a total of eight variant-gene pairs were identified where the allele associated with protection against cALT was also associated with reduced transcription levels. Furthermore, sQTL analysis in GTEx v8 identified two genes in the liver (*HSD17B13* and *ANPEP)* and 13 genes that were affected in at least two tissues (**Supplemental Table 24**). Finally, two of our lead SNPs were in high LD (r^2^ > 0.8) with variants that regulated H3K27ac levels in liver tissue (hQTLs), namely *EFHD1* (hQTL SNPs rs2140773, rs7604422 in *EFHD1*) and *FADS2* loci (hQTL rs174566 in *FADS2*)^44^.

#### Assay for chromatin accessibility using liver-derived cells

We next mapped our cALT loci to regions of open chromatin using ATAC-seq in three biologically-relevant liver-derived tissues (human liver, liver cancer cell line [HepG2], and hepatocyte-like cells [HLC] derived from pluripotent stem cells)^45^. Additionally, we used promoter-focused Capture-C data to identify those credible sets that physically interact with genes in two relevant cell types (HepG2 and liver). For each credible set, we identified genes with significant interactions (CHiCAGO score > 5, **Methods**) that overlap with at least one lead variant (**Supplemental Table 25**). These datasets are useful entry points for deciphering regulatory mechanisms involved in the pathophysiology of NAFLD. Based on DEPICT gene prediction, coding variant linkage analysis, and QTL colocalization (**Supplemental Tables 18-25**), 215 potentially relevant genes were identified for the 77 loci. A protein-protein interaction (PPI) analysis revealed that among the 192 available proteins, 86 nodes were observed, with strong PPI enrichment (P < 9.0×10^-8^) indicating that the protein network shows substantially more interactions than expected by chance (**Supplemental Table 26 and Supplemental Figure 12**).

### Variant-to-Gene Ensembl Mapping Approach to nominate putative causal genes

We developed an ensemble method for predicting the likely causal effector gene at 77 loci based on 8 distinct gene-mapping analyses, including: SNP-gene overlap, DEPICT gene prediction, coding variant linkage, colocalization with eQTL, sQTL and hQTL, promotor Capture-C and/or ATAC-Seq peak overlap, and PPI network analysis. For each gene that resides in a sentinel locus, the number of times that it was identified in the eight analyses was summarized into a nomination score which reflects the cumulative evidence that the respective gene is the causal effector gene in the region. This ensemble method for mapping variants to genes resulted in the nomination of a single gene as the causal effector gene at 53 of 77 genomic loci. At the remaining 24 loci, two loci lacked any data to support the nomination of a causal gene, and at 22 loci two or more causal genes were nominated because they shared the maximum nomination score (**Supplemental Table 27**). We highighted 35 loci for which a causal gene was prioritized by at least 3 sources of evidence (or 4 sources of evidence for coding variants) in **Table 1a/b.** These included 6 loci with co-localizing eQTLs in liver or adipose tissues and connection to the predicted gene via Promoter CaptureC data (i.e., *EPHA2, IL1RN, SHROOM3, HKDC1, PANX1, DHODH;HP)*.

**Table 1a.**
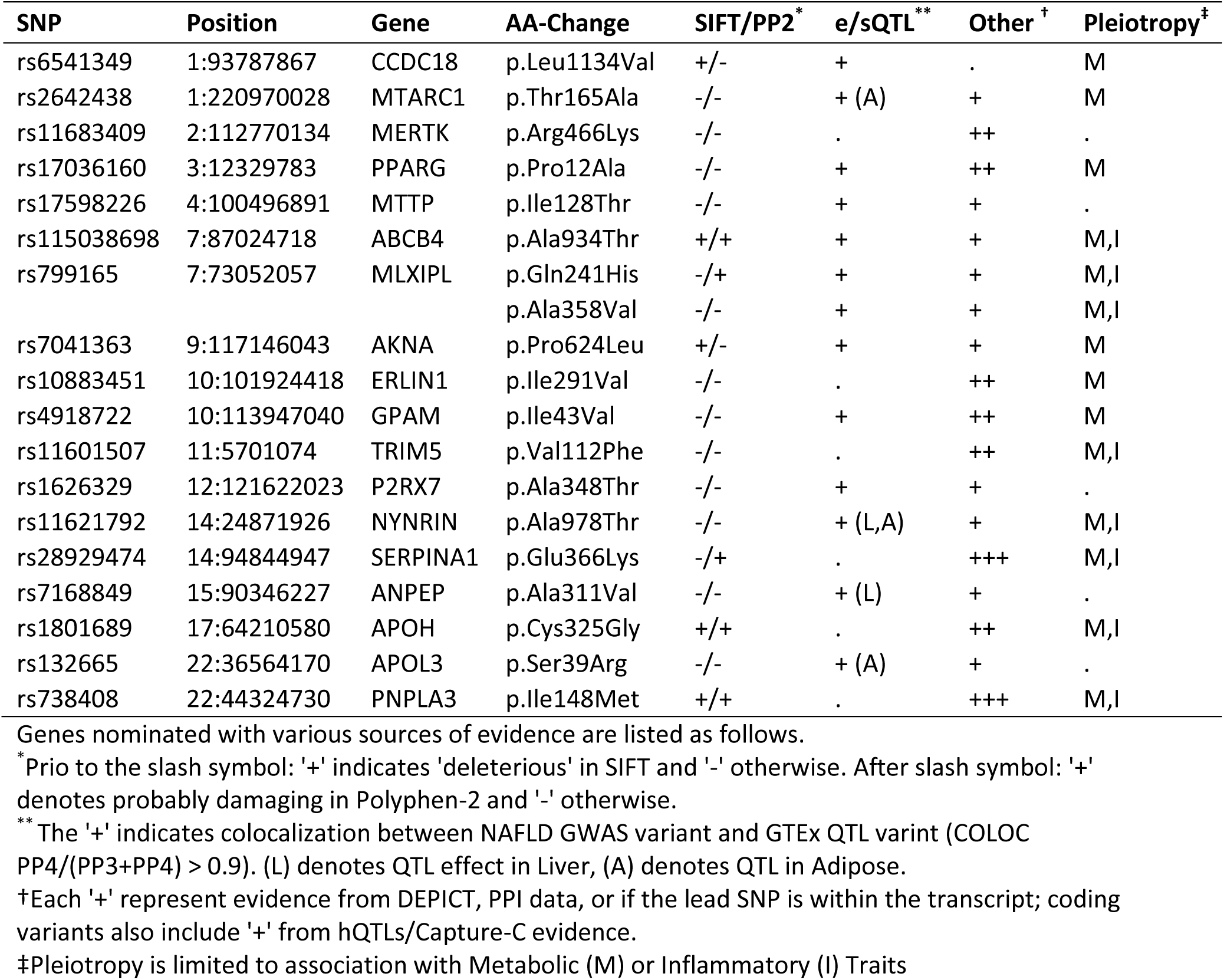
Gene nominations at loci with strongest evidence for coding variants.

**Table 1b.** Gene nominations at loci with strongest evidence for non-coding variants.

| SNP | Position | Gene | hQTL | CaptureC | e/sQTL** | Other<sup>†</sup> | Pleiotropy<sup>‡</sup> |
| --- | --- | --- | --- | --- | --- | --- | --- |
| rs36086195 | 1:16510894 | EPHA2 | . | + | + (L,A) | + | M |
| rs6734238 | 2:113841030 | IL1RN | . | + | + (A) | ++ | I |
| rs10201587 | 2:202202791 | CASP8 | . | + | + | + | M |
| rs11683367 | 2:233510011 | EFHD1 | + | . | + (L) | + | . |
| rs61791108 | 3:170732742 | SLC2A2 | . | + | . | +++ | M |
| rs7653249 | 3:136005792 | PCCB | . | . | + | ++ | M,I |
| rs12500824 | 4:77416627 | SHROOM3 | . | + | + (L) | + | M |
| rs10433937 | 4:88230100 | HSD17B13 | . | . | + (L,A) | + | M,I |
| rs799165 | 7:73052057 | BCL7B | . | + | + | + | M,I |
| rs687621 | 9:136137065 | ABO | . | . | + | + | M,I |
| rs35199395 | 10:70983936 | HKDC1 | . | + | + (L,A) | + | M |
| rs174535 | 11:61551356 | FADS2 | + | . | + (A) | ++ | M,I |
| rs56175344 | 11:93864393 | PANX1 | . | . | + (L,A) | ++ | . |
| rs34123446 | 12:122511238 | MLXIP | . | + | + | + | M,I |
| rs12149380 | 16:72043546 | DHODH | . | + | + | + | M,I |
|  |  | HP | . | + | + (A) | . | M,I |
| rs2727324 | 17:61922102 | DDX42 | . | + | + | + | M |
|  |  | SMARCD2 | . | . | + | + | M |
| rs5117 | 19:45418790 | APOC1 | . | . | + | ++ | M,I |

Genes nominated with various sources of evidence are listed as follows.
\*Prio to the slash symbol: '+' indicates 'deleterious' in SIFT and '-' otherwise. After slash symbol: '+' denotes probably damaging in Polyphen-2 and '-' otherwise.
\*\* The '+' indicates colocalization between NAFLD GWAS variant and GTEx QTL variant (COLOC PP4/(PP3+PP4) > 0.9). (L) denotes QTL effect in Liver, (A) denotes QTL in Adipose.
†Each '+' represent evidence from DEPICT, PPI data, or if the lead SNP is within the transcript; coding variants also include '+' from hQTLs/Capture-C evidence.
‡Pleiotropy is limited to association with Metabolic (M) or Inflammatory (I) Traits

#### Gene expression of nominated genes in the Liver Single Cell Atlas

To confirm that the nominated genes are involved in liver biology, we performed a gene expression lookup in single cell RNA-Seq data from the Liver Single Cell Atlas^46^. As a result, for 76 of 77 loci a gene was nominated that was expressed in at least one liver cell type, with exception of the rs9668670 locus which nominated several keratin genes (*KRT84;KRT82;KRT74*)(**Supplemental Table 27**).

### Transcription factor analysis

We observed that 14 nominated genes are transcription factors (TF) (**Supplemental Table 28**). Using the DoRothEA data in OmniPath, we identified that two of these TFs have several downstream target genes that were also identified in our GWAS scan (**Methods**). Notably, the CEBPA TF targets the downstream genes *PPARG*, *TRIB1*, *GPAM*, *FTO*, *IRS1*, *CRIM1*, *HP, TBC1D8*, and *CPS1*, but also *NCEH1,* a gene in the vicinity of one of our associations that lacked a nominated candidate gene. Similarly, *HNF1A*, the lead gene in the EA scan (and corresponding to the trans-ancestry *P2RX7* locus) targets *SLC2A2*, *MTTP*, and *APOH*.

### Pleiotropy and related-trait genetic architecture of lead cALT SNPs

We next sought to identify additional traits that were associated with our 77 trans-ancestry lead SNPs using four different approaches. First, a LabWAS of distinct clinical laboratory test results^47^ in MVP (**Methods**) yielded 304 significant SNP-trait associations (adjusted Bonferroni nominal P = 3.1×10^-5^, **Supplemental Table 29, Supplemental Figure 13**). Second, a PheWAS Analysis in UK Biobank data using SAIGE (**Methods**) identified various SNP-trait associations that mapped to loci previously associated with liver and cardiometabolic traits, as well as additional enriched association for gallstones, gout, arthritis, and hernias (adjusted Bonferroni nominal P < 4.6×10^-7^**, Supplemental Table 30**). In particular, we examined all associations for PheCode 571.5, “Other chronic nonalcoholic liver disease” which comprised 1,664 cases and 400,055 controls, which with a disease prevalence of 0.4% seems to be underreported. Still, of the 73 variants with available data, 14 were both nominally associated and directionally consistent with our scan (signed binomial test P=3.4×10^-9^), providing additional validation for our scan (**Supplementary Table 31**). Third, a SNP lookup using the curated data in the MRC IEU OpenGWAS project (**Supplemental Table 32**) identified 2,892 genome-wide significant SNP-trait associations for trans-ancestry SNPs, with additional 283 SNP-trait associations for the ancestry-specific lead SNPs. Finally, we performed cross-trait regional colocalization analyses of EA, AA, and HISP lead loci with 36 other GWAS statistics of cardiometabolic and blood cell related traits (**Methods**). This resulted in significant regional colocalization for 64 SNP-trait pairs in EA, 32 SNP-trait pairs in AA, and 12 SNP trait pairs in HISP (**Supplemental Table 33**).

Based on the four analyses described above, we selected all SNP-trait associations relevant phenotypes to NAFLD biology and classified them as liver (e.g. ALT, ALP, AST, and GGT), metabolic (e.g. HDL, LDL, and total cholesterol, triglycerides, BMI, glucose, and HbA1c), or inflammatory traits (e.g., C-reactive protein, white blood cell count, lymphocyte count, granulocytes, neutrophils, monocyte count, basophils, eosinophils, and myeloid white cells) (**Supplemental Tables 29-33 Figure 3**). Across the trans-ancestry lead variants (n=77), ancestry-specific variants (n=3), and secondary proximal associations (*HNF1A,* n=1), 17 trans-ancestry and one EA-specific SNPs showed association with only liver traits (**Figure 3).** In contrast, 17 trans-ancestry and 3 ancestry-specific loci showed associations with both liver and metabolic traits whereas 4 trans-ancestry loci showed associations with both liver and inflammatory traits. Finally, 39 trans-ancestry loci showed association with all three traits: liver enzymes, cardiometabolic traits, and inflammation, including 15 of 17 loci that were externally validated in liver biopsy and/or imaging cohorts (color-coded in red in **Figure 3**). Collectively, our findings identify novel cALT-associated genetic loci with pleotropic effects that may impact hepatic, metabolic and inflammatory traits.

**Figure 3.**
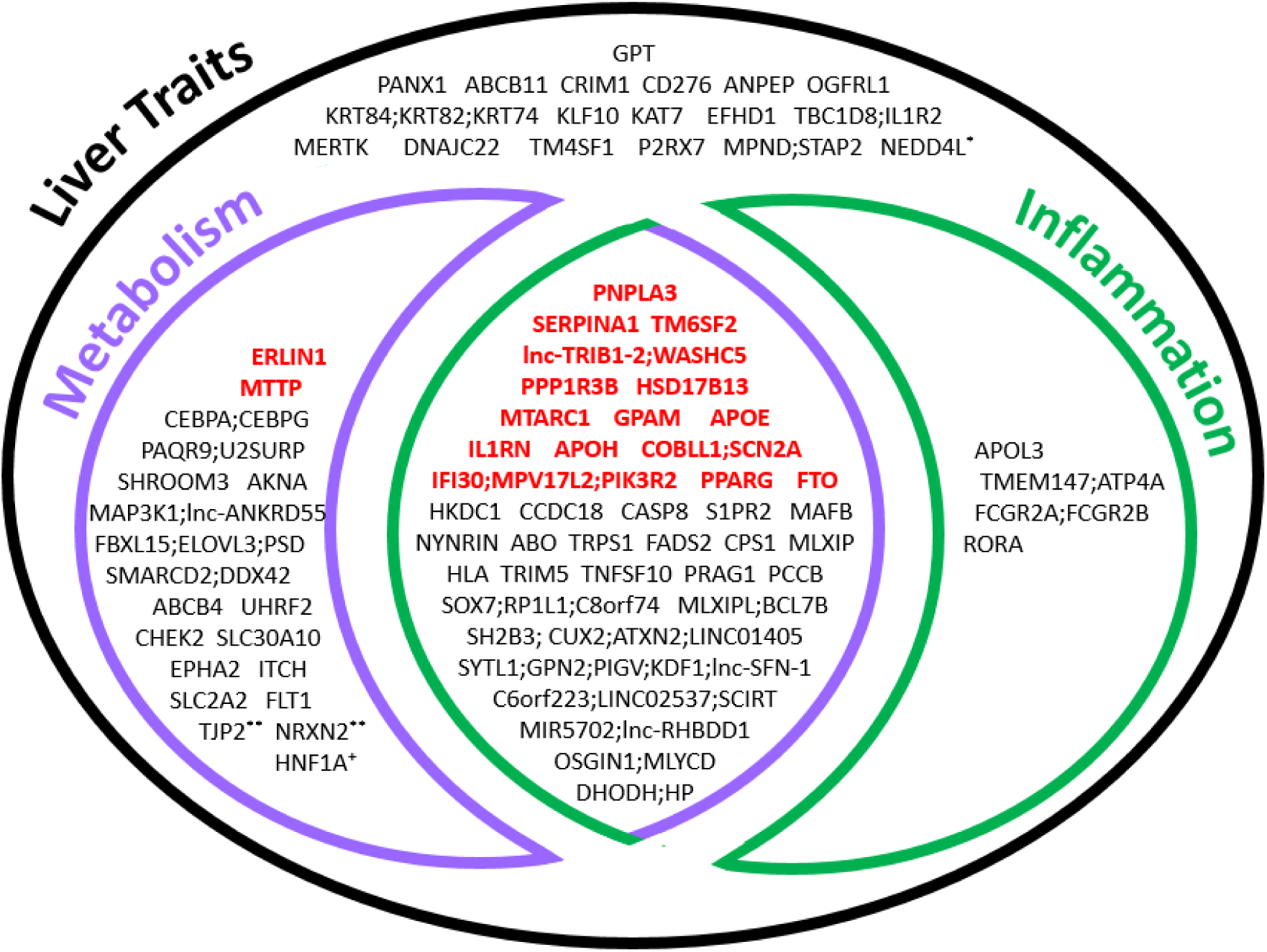
Venn diagram depicting overlapping liver, metabolic and inflammatory traits among NAFLD-associated loci. Overlapping liver (blackblack), metabolic (purplepurple) and/or inflammatory (green) traits are shown in association with 77 trans-ancestry and additional ancestry-specific lead SNPs. The trait categorizations reflect significant SNP-trait associations identified by: 1) LabWAS of clinical laboratory results in MVP; 2) PheWAS with UKBB data using SAIGE; 3) SNP lookup using the curated data in the IEU OpenGWAS projects; and 4) cross-trait colocalization analyses using COLOC of EA, AA and HISP lead loci with 36 other GWAS statistics of cardiometabolic and blood cell related traits. GenesGenes denoted in bold and color-coded in red refer to the loci also associated with quantitative hepatic fat on imaging analyses or histologically characterized NAFLD from liver biopsies. *Locus identified in European-only GWAS. ** Locus identified in African American-restricted analysis. ⁺Secondary signal from European analysis (e.g. HNF1A/P2RX7).

### Pleiotropy-stratified GRS and histological NAFLD

The foregoing analyses raised the possibility that SNPs with greater pleiotropy relative to metabolic and/or inflammatory traits beyond liver-related traits may have greater contributions to NAFLD. To this end, we compared the GRS between four subgroups of trans-ancestry SNPs as defined in **Figure 3** Venn Diagram including: (i) 17 SNPs only associated with liver traits; (ii) 5 SNPs associated with liver and inflammatory traits; (iii) 17 SNPs associated with liver and metabolic traits; and (iv) 38 SNPs associated with liver, cardiometabolic and inflammatory traits. As shown in **Supplemental Table 9D**, all 4 subgroups showed significant capacity to predict NAFLD. However, the strongest effect was observed for the group combining all three traits (Beta = 0.592, P=2.8 x 10^-9^) followed by liver and metabolic traits (Beta = 0.148, P=4.6 x 10^-6^), whereas liver trait alone and liver with inflammatory traits showed significant but reduced effects **(Supplementary Figure 14)**. Collectively, these findings show greater NAFLD associations for pleiotropic SNPs that combine liver, metabolic and inflammatory traits.

### Directional Pleiotropy and Associated Gene Clusters

Finally, we visualized the direction and strength of the associations between the 77 trans-ancestry loci and 7 inflammatory biomarkers and 13 cardiometabolic traits in a heatmap **(Figure 4).** The loci were grouped into 7 broad gene clusters using stratified agglomerative hierarchical clustering **(Methods)**. Gene cluster 1 consisted of 5 trans-ethnic loci (including *APOE*), for which cALT risk alleles were associated with increased LDL and total cholesterol, apolipoprotein B1, and markers of inflammation. Gene cluster 2 comprised of genes (such as *IL1RN*, *MTARC1*, *GPAM*, and *TRIB1*) for which the cALT risk alleles were associated with increased LDL and total cholesterol, apolipoprotein B1, but decreased levels of inflammatory markers. The majority of the SNPs in cluster 1 and 2 were additionally characterized by lower triglyceride levels, but not all. Gene cluster 3 (including *MTTP*) included genes that showed predominantly positive associations with apolipoprotein B1, LDL and total cholesterol. Gene cluster 4 was characterized by a lack of distinctive biomarker co-association profiles. The genes in cluster 5 (including *PNPLA3*, *ERLIN1*, and *PPP1R3B*) were characterized by higher rates of type 2 diabetes, but decreased levels of triglycerides, LDL cholesterol, HDL cholesterol, apolipoprotein B1, apolipoprotein A1, white blood cell count, and neutrophils. The genes in cluster 6 (e.g., *PPARG*, and *SLC30A10*) were associated with higher triglycerides and type 2 diabetes, but decreased sex hormone binding globulin (SHBG), HDL cholesterol, and apolipoprotein 1A. Finally, the 3 genes in cluster 7 (including *TM6SF2* and *FTO*) were associated with increased inflammatory markers, but lower apolipoprotein B1, total and LDL cholesterol. Interestingly, for a total of 9 SNPs (*TRIB1*, *PPARG*, *SLC30A10* [former *LYPLAL1*], *MLXIP*, *CEBPA*, *COBLL1*, *C6orf223*, *MIR5702*, and *SH2B3*) the cALT risk allele was associated with lower BMI, consistent with a “lean NAFLD” phenotype. Similarly, the cALT risk alleles of *SERPINA1* and *OSGIN1* loci seemed to be associated with lower rates of type 2 diabetes, and *SH2B3* and *SLC2A2* with lower glucose and HbA1c. Overall, these directional associations define distinct characteristics for each loci and clusters with potential biological implications.

**Figure 4:**
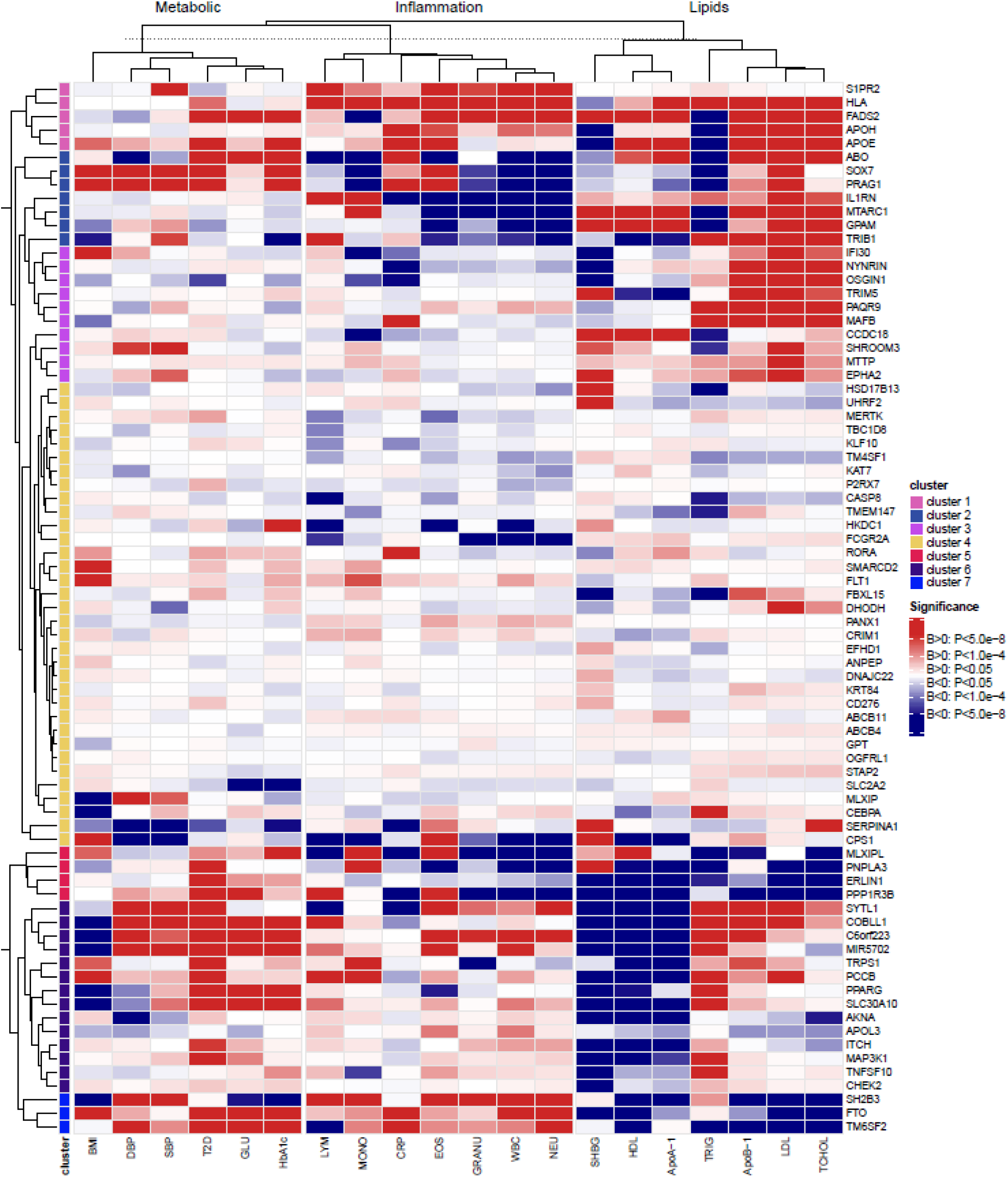
Seven gene clusters with distinct biomarker association profiles. The 77 loci cluster along 7 groups using stratified linkage hierarchical clustering. Each cluster has a distinct biomarker association profile, which is visualized with a heatmap. Twenty traits are clustered within their biological strata (e.g. lipids, inflammation, and metabolic). The color coding corresponds to the direction of association of the cALT risk allele (red, positive association; blue, negative association) and the strength of the association based on the P-value.

## Discussion

In this study, we describe the first of its kind multi-ancestry GWAS of cALT as a proxy for NAFLD, which resulted in a total of 77 trans-ancestry loci, of which 25 have not been associated with NAFLD or ALT before. We additionally identified three ancestry-specific loci, as well as 29 conditionally independent SNPs in EAs and six in AAs. We assembled two external replication cohorts with histologically confirmed NAFLD (7,397 NAFLD cases and 56,785 population controls) and hepatic fat defined by imaging (n = 44,289), and validated the association of 17 SNPs with NAFLD, of which nine are novel (*TRIB1*, *PPARG*, *MTTP*, *SERPINA1*, *FTO*, *IL1RN*, *COBLL1*, *APOH*, and *IFI30*). Furthermore, a GRS based on novel SNPs alone was predictive of histologically defined NAFLD.

Pleiotropy analysis allowed us to characterize the genetic architecture of NAFLD and all validated SNPs showed significant associations with metabolic risk factors and/or inflammatory traits, the most common being plasma lipid-related, followed by glycemic traits, hypertension, and cardiovascular disease. A GRS based on the subset of SNPs that are associated with cALT, cardiometabolic traits, and inflammatory markers showed the highest capacity to predict histological NAFLD. Our ensemble variant-to-gene mapping method nominated a single causal effector gene at 53 genomic loci. We found that these genes were highly expressed in one or more cell types in the liver and have prior biological connections to liver metabolism, physiology, or disease, making this list compelling for further interrogation. Our directional pleiotropy analysis for metabolic risk factors are overall concordant with the results from Sliz et al, which investigated 4 NAFLD SNPs (*LYPLAL1*, *PNPLA3*, *GCKR*, and *TM6SF2*).^48^ In addition to associations with inflammatory markers, we show that the risk alleles of *TM6SF2*, *PNPLA3*, and *SLC30A10* (former *LYPLAL1*) are positively associated with type 2 diabetes and HbA1c. Collectively, our findings offer a comprehensive, expanded, and refined view of the genetic contribution to cALT with potential clinical, pathogenic, and therapeutic relevance.

Our proxy NAFLD phenotype was based on chronic ALT elevation with the exclusion of other known diagnoses of liver disease or causes of ALT elevation (e.g. viral hepatitis, alcoholic liver disease, hemochromatosis), based on previous validation within VA population^21, 31^. In this regard, several GWAS studies of liver enzyme levels have been reported, particularly of serum ALT^10, 11, 16, 34, 35^, but not all studies systematically excluded other causes of ALT elevation. For example, Pazoki *et al*. recently reported 230 locis related to ALT, of which 52 were also included in our panel of 77 (67.5%) lead cALT loci^34, 35^. While our cALT approach was designed to enhance the specifity of non-invasive NAFLD diagnosis^21, 31^, this overlap is not surprising given the high prevalence of NAFLD in the general population. Furthermore, we noted that inflammatory traits were associated with over half of the cALT loci in our study. Indeed, while ALT can be normal in patients with hepatic steatosis, we focused on chronic ALT elevation that represents ongoing hepatocellular injury that can promote liver disease progression. For our controls, we excluded individuals with mild ALT elevation and selected healthier ‘super-controls’ to minimize potential phenotype misclassification (e.g. attributing cases as controls). We recognize that some cALT loci such as GPT may be involved more directly in ALT biology rather than NAFLD. Based on the observation that replicated SNPs had higher effect sizes in the histological cohort than for cALT, we hypothesize that 22 non-replicated SNPs with higher effect sizes for histological NAFLD than cALT are candidate loci for NAFLD. In addition, our pleiotropy analyses suggest that 24 non-replicated cALT SNPs associated with metabolic and inflammatory traits are more likely to be related to NAFLD, of which 12 SNPs overlap with the list of candidate NAFLD loci.

The MVP is one of the world’s largest and most ancestrally diverse biobanks, of which 25% of the participants are of non-white ancestry, and this diversity enhanced the value of this study. Utilizing data from multiple ancestries allowed us to narrow down putative causal variants for NAFLD through trans-ancestry fine-mapping. Moreover, we identified two cALT NAFLD loci specifically in AAs. For example, the lead SNP at the *ABCB4* locus (rs115038698) was in high LD with the missense variant rs61730509 (Ala934Thr, AFR r^2^=0.92) and had a very potent effect (OR=1.87, CI=1.64-2.14, P=1.8×10^-20^). This variant is of low frequency in AA (MAF=1.2%) but virtually absent in EA and ASN. *ABCB4*, also known as multidrug resistance protein 3 (*MDR3*), is a compelling candidate gene, as it is involved in hepatocyte lipid transport and has been previously implicated in cholestasis, gallbladder disease, and adult biliary fibrosis/cirrhosis^49–51^.

Ultimately, the affirmative external validation of our lead cALT loci in NAFLD cohorts with liver biopsies and imaging supports the relevance of our proxy phenotype for NAFLD. A total of 17 loci associated with cALT were also significantly associated with hepatic fat based on liver biopsy and/or radiological imaging. These included loci previously associated with NAFLD or all-cause cirrhosis (e.g., *PNPLA3, TM6SF2, HSD17B13, MTARC1, ERLIN1, GPAM, and APOE*), but also included several of the novel loci reported here (e.g., *TRIB1, SERPINA1, MTTP, IL1RN, IFI30, COBLL1, APOH, FTO, PPP1R3B and PPARG*). For all loci except *PPP1R3B,* we observed concordant directionality of effects between cALT and hepatic fat. The apparent discrepancy in the *PPP1R3B* locus has been reported before^12^ and may represent diffuse attenuation on radiologic images due to hepatic accumulation of glycogen^36, 52^ rather than triglycerides^53^. These novel and validated genes make excellent gene candidates for NAFLD. In addition, our study failed to replicate the GCKR locus, where a common missense variant (rs780094) has been repeatedly shown to confer susceptibility to NAFLD ^20^. The SNP is a risk factor for increased triglycerides, C-reactive protein, LDL cholesterol, but seems to be protective for T2D, fasting glucose, alcohol intake, AUD, BMI, and monocyte percentage. It his hypothesized that the variant GCKR protein loses interaction efficiency with glucokinase, which promotes hepatic glucose metabolism, decreases plasma glucose levels, and increases NAFLD risk^54^. Our phenotype might not be a suitable proxy for NAFLD for SNPs that act through multiple pathways with opposing effects on ALT.

A substantial fraction of our cALT loci showed a shared genetic co-architecture with metabolic traits (**Figure 4**). Of interest is that for 9 SNPs the cALT risk allele was associated with lower BMI, including PPARG. These SNPs seem to exhibit mild lipodystrophic effects, characterized by reduced adipose tissue and increased hepatic steatosis. Further study is required to clarify whether and which loci are working primarily in adipose tissue with a secondary effect on liver steatosis.

Several genes and liver-enriched transcription factors involved in LDL and triglyceride pathways have been indentified, such as the liver-biopsy and/or imaging validated *TRIB1, FTO*^55, 56^*, COBLL1*^57, 58^*, MTTP, TM6SF2, PPARG, APOE,* and *GPAM*^59–62^, but also the cALT-only associated variants in *MLXIPL, MLXIP, CEBPA, FADS2, APOH, RORA, and HNF1A*. *TRIB1* presumably regulates VLDL secretion by promoting the degradation carbohydrate-response element binding protein (ChREBP, encoded by *MLXIPL*), reducing hepatic lipogenesis and limiting triglyceride availability for apolipoprotein B (apoB) lipidation. Furthermore, *TRIB1* co-activates the transcription of *MTTP*, which encodes the microsomal triglyceride transfer protein that loads lipids onto assembling VLDL particles and facilitates their secretion by hepatocytes. Lomitapide, a small molecule inhibitor of *MTTP*, is approved as a treatment for lowering LDL cholesterol in homozygous familial hypercholesterolemia^63^ and a potential therapeutic target for NAFLD. *TRIB1* is also involved in the degradation of the key hepatocyte transcription factor, CCAAT/enhancer-binding protein alpha (*CEBPA*)^64^, which together with *HNF1A* (HNF1 Homeobox A), *RORα* (retinoic acid receptor-related orphan receptor-α) and *MIR-122* are involved in a feedback loop of the the liver-enriched transcription factor network to control hepatocyte differentiation^65^. RORα is also a suppressor of transcriptional activity of peroxisome proliferator-activated receptor γ (PPARγ)^66, 67^. *PPARG*, encoding PPARγ, upregulates LDL-receptor-related protein 1 (*LRP1*), which facilitates the hepatic uptake of triglyceride-rich lipoproteins via interaction with apolipoprotein E (apoE)^68, 69^. *PPARG* is predominantly expressed in adipose tissue, and hepatic expression levels of PPARG are significantly increased in patients with NAFLD^70^. Large randomized controlled clinical trials have reported that the *PPARG* agonists rosiglitazone and pioglitazone improve NAFLD-related hepatic steatosis, hepatic inflammation, and fibrosis^71–75^. However, the treatment is frequently accompanied with weight gain and fluid retention, limiting its application and potential long-term drug adherence. RORα however, competes with PPARγ for binding to PPARγ target promoters, and therapeutic strategies designed to modulate RORα activity in conjunction with PPARγ may be beneficial for the treatment of NAFLD. ApoE and ApoH^59^ play an important role in the production and clearance of VLDL by facilitating the hepatic uptake of triglyceride-rich lipoproteins^76–79^. ApoE deficiency is suggested to affect hepatic lipid deposition in dietary-challenged murine models^80^. Similarly, a Western high-fat cholesterol-rich diet accelerates the formation of NASH with fibrosis in ApoE-deficient mice^81^.

More than half of our cALT loci had a significant association with at least one inflammatory trait (**Figure 4**), consistent with the multiple-hit hypothesis of NAFLD^82^. For example, the transcription factor MafB regulates macrophage differentiation^83^ and genetic variation in *MAFB* has been associated with hyperlipidemia and hypercholesterolemia^27^. Mice with macrophage-specific Mafb-deficiency are more susceptible to obesity and atherosclerosis^84, 85^. *FADS1* and *FADS2* are markedly induced during monocyte to macrophage differentiation, and it is hypothesized that they impact metabolic disease by balancing proinflammatory and proresolving lipid mediators^86, 87^. Another interesting locus is *IL1RN*, which in our study is associated with lower risk of cALT and liver-biopsy characterized NAFLD. GTEx data shows heterogeneous directions of effect across various tissues, with the minor G-allele being associated with increased *IL1RN* expression in liver but decreased expression in subcutaneous adipose tissue. *IL1RN* encodes the anti-inflammatory cytokine interleukin-1 receptor antagonist (IL-1Ra) and is a natural inhibitor of IL-1 activity by blocking the binding of IL-1β to IL-1R, and is considered a potential therapeutic target for NAFLD treatment^88^. IL-1β has been shown to lead to chronic low-grade inflammation^89–91^, insulin resistance and hepatic fat, and promotes hepatic steatosis, inflammation and fibrosis^92, 93^. A study in mice has shown that IL-1R-deficiency protects from liver fibrosis^94^, and the deletion of IL-1R reduces liver injury in acute liver disease by blocking IL-1 driven autoinflammation^95^. In two studies of patients with diabetes, blockade of IL-1 signaling with Anakinra (a recombinant form of IL-1Ra) improved glycemic control^96–98^. It remains to be investigated whether remodeling of the adipose tissue inflammasome via IL-1 signaling blockade in obesity-associated NAFLD offers potential therapeutic benefit. Other loci implicated with inflammatory traits include *RORA*^99, 100^, *IFI30*^101, 102^, *CD276*^103–105^, *FCGR2A*^106^, and *P2RX7*^107–109^. Interestingly, these inflammation-related genes and pathways emerged from our cALT GWAS despite the indirect assessment of our phenotype, clearly implicating inflammation early in NAFLD.

Finally, *PANX1* and *MERTK* genes that are associated with liver traits only seem particulary interesting. For *PANX1*, the directional concordance of effects between cALT and *PANX1* gene expression in the liver suggests possible relevance as a therapeutic target. It has been reported that the genetic deletion of Pannexin 1 (encoded by *PANX1*) has a protective effect in a mouse model of acute and chronic liver disease^110, 111^, and our data demonstrate that a SNP near *PANX1* was associated with reduced *PANX1* expression and reduced risk of NAFLD. These data nominate *PANX1* as a therapeutic target for silencing in NAFLD treatment. For MerTK, two missense variants (Arg466Lys and Ile518Val, r^2^=0.98) were predicted to affect protein function. MerTK signaling in hepatic macrophages was recently shown to mediate hepatic stellate cell activation and promote hepatic fibrosis progression^112^. Variants in *MERTK* were associated with liver fibrosis progression in HCV-infected patients^113^, raising the possibility for MerTK as a novel therapeutic target against fibrosis^114^. We emphasize that functional studies of our nominated causal genes are needed to demonstrate casual relevance, their impact on hepatosteatosis, and ultimately to determine their underlying mechanisms.

In conclusion, we discovered 77 genomic loci associated with cALT in a large, ancestrally diverse cohort. Our cases of cALT were excluded for other known causes of liver disease or elevated ALT and, not surprisingly, were substantially enriched for metabolic disorders. We replicated our findings in external cohorts with hepatic fat defined by liver biopsy or radiologic imaging. The genetic architecture of the lead loci indicate a predominant involvement of metabolic and inflammatory pathways. This study constitutes a much-needed large-scale, multi-ancestry genetic resource that can be used to build genetic prediction models, identify causal mechanisms, and understand biological pathways contributing to NAFLD initiation and disease progression.

## Methods

We performed a large-scale trans-ancestry NAFLD GWAS in the Million Veteran Program. We subsequently conducted analyses to facilitate the prioritization of these individual findings, including transcriptome-wide predicted gene expression, secondary signal analysis, coding variant mapping, variant-to-gene mapping, and pleiotropy analysis to fine-map the genomic loci to putatively causal genes and biological mechanisms.

### Discovery cohort in Million Veteran Program

The Million Veteran Program (MVP) is a mega-biobank that was launched in 2011 and supported entirely by the Veterans Health Administration (VA) Office of Research and Development in the United States (US) of America, to develop a genetic repository of US Veterans with additional information through the VA electronic health record system and MVP questionnaires to learn how genes, lifestyle and military exposure affect health and disease. The MVP received ethical and study protocol approval from the VA Central Institutional Review Board (IRB) in accordance with the principles outlined in the Declaration of Helsinki. Over 60 VA Medical Centers have participated in this study nationally. The specific design, initial demographics of the MVP have been detailed previously^29^. Electronic health record information from the VA’s Corporate Data Warehouse (CDW) was used for clinical and demographic information. For genetic analyses, DNA extracted from whole blood was genotyped in customized Affymetrix Axiom Array which contains a total of 723,305 SNPs enriched for: 1) low frequency variants in AA and HISP populations, and 2) variants associated with diseases common to the VA population^29^. Further quality control procedures have been previously described^115^.

#### Proxy NAFLD Phenotype

MVP NAFLD phenotype definitions were developed by combining a previously published VA CDW ALT-based approach with non-invasive clinical parameters available to practicing clinicians at the point of care ^21, 31^. The primary NAFLD phenotype (labeled “ALT-threshold”) was defined by: (i) elevated ALT >40 U/L for men or >30 U/L for women during at least two time points at least 6 months apart within a two-year window period at any point prior to enrollment and (ii) exclusion of other causes of liver disease (e.g., presence of chronic viral hepatitis B or C (defined as positive hepatitis C RNA > 0 international units/mL or positive hepatitis B surface antigen), chronic liver diseases or systemic conditions (e.g., hemochromatosis, primary biliary cholangitis, primary sclerosing cholangitis, autoimmune hepatitis, alpha-1-antitrypsin deficiency, sarcoidosis, metastatic liver cancer, secondary biliary cirrhosis, Wilson’s disease), and/or alcohol use disorder (e.g., alcohol use disorder, alcoholic liver disease, alcoholic hepatitis and/or ascites, alcoholic fibrosis and sclerosis of liver, alcoholic cirrhosis of liver and/or ascites, alcoholic hepatic failure and/or coma, and unspecified alcoholic liver disease). The control group was defined by having a: normal ALT (≤30 U/L for men, ≤20 U/L for women) and no apparent causes of liver disease or alcohol use disorder or related conditions^21^. Habitual alcohol consumption was assessed with the age-adjusted Alcohol Use Disorders Identification Test (AUDIT-C) score, a validated questionnaire annually administered by VA primary care practitioners and used previously in MVP^116, 117^. Demographics of the NAFLD cohort are shown in **Supplementary Table 1**. The prevalence of cirrhosis and advanced fibrosis was based on the following ICD-9 codes: 456.2, 456.21, 571.5, 572.2, 572.3, and ICD-10 codes: K72.9, K72.91, K74.0, K74.02, K74.1, K74.2, K74.6, K74.69

### Single-variant autosomal analyses

We tested imputed SNPs that passed quality control (i.e. HWE > 1×10^-10^, INFO > 0.3, call rate > 0.975) for association with NAFLD through logistic regression assuming an additive model of variants with MAF > 0.1% in European American (EA), and MAF > 1% in African Americans (AA), Hispanic Americans (HISP), and Asian Americans (ASN) using PLINK2a software^118^. Indels were excluded from analysis. Covariates included age, gender, age-adjusted AUDIT-C score, and first 10 principal components (PC’s) of genetic ancestry. We aggregated association summary statistics from the ancestry-specific analyses and performed a trans-ancestry meta-analysis. The association summary statistics for each analysis were meta-analyzed in a fixed-effects model using METAL with inverse-variance weighting of log odds ratios^119^. Variants were clumped using a range of 500kb and/or CEU r^2^ LD > 0.05, and were considered genome-wide significant if they passed the conventional p-value threshold of 5×10^-8^. Trans-ancestry and ancestry-specific summary statistics are displayed in **Supplementary Tables 2-5.**

### Secondary signal analysis

The PLINK --condition and --condition-list parameters were used to conduct stepwise conditional analyses on individual level data in MVP to detect ancestry-specific distinct association signals nearby lead SNPs. Regional SNPs were eligible if they were located within 500kb of lead SNP, had a MAF >1% and passed standard quality control criteria (INFO > 0.3, HWE P > 1.0×10^-10^, call rate > 0.975). Logistic regression was performed in a stepwise fashion, starting with a regional association analysis with the following set of covariates: lead SNP imputed allele dosage, age, gender, and 10 PC’s of genetic ancestry. If the corresponding output file contained SNP(s) that reached locus-wide significance (P < 1.0×10^-5^), the most significant SNP was selected and added to the covariate set. The regression was repeated until no locus-wide significant SNPs remained. Secondary signals are shown in **Supplementary Table 11.**

### Credible Sets

We calculated Wakefield’s approximate Bayes’ factors^41^ based on the marginal summary statistics of the trans-ancestry meta-analysis and ancestry specific summary statistics using the CRAN R package corrcoverage^120^. For each locus, the posterior probabilities of each variant being causal were calculated and a 95% credible set was generated which contains the minimum set of variants that jointly have at least 95% posterior probability (PP) of including the causal variant **(Supplementary Tables 12-15)**.

### External validation in a Liver Imaging cohort

A replication lookup of lead loci was performed to evaluate the extent to which genetic predictors of hepatocellular injury (cALT) correspond with quantitative hepatic fat derived from computed tomography (CT) / magnetic resonance imaging (MRI)-measured hepatic fat in the Penn Medicine Biobank (PMBB), UK Biobank, Multi-Ethnic Study of Atherosclerosis (MESA), Framingham Heart Study (FHS), and University of Maryland Older Order Amish study **(Supplementary Table 8)**. Attenuation was measured in Hounsfield units. The difference between the spleen and liver attenuation was measured for PMBB; a ratio between liver attenuation/spleen attenuation was used for MESA and Amish; and liver attenuation/phantom attenuation ratio in FHS as previously described by Speliotes *et al*^12^. Abdominal MRI data from UK Biobank data were used to quantify liver fat using a two-stage machine learning approach with deep convolutional neural networks^121^. CT-measured hepatic fat was estimated using a multi-stage series of neural networks for presence of scan contrast and liver segmentation using convolutional neural networks. The PMBB included CT data on 2,979 EA and 1,250 AA participants^122^, the FHS included a total of 3,011 EA participants, the Amish study 754 EA participants, and MESA contributed 1,525 EA, 1,048 AA, 923 HISP, and 360 ASN participants for concordance analysis. The UK Biobank included MRI image data from 36,703 EA participants. All cohorts underwent individual-level linear regression analysis on hepatic fat, adjusted for the covariates of age, gender, first 10 principal components of genetic ancestry, and alcohol intake if available. If the lead SNP was not available in any of the studies, a proxy SNP in high LD with the lead variant was used (r^2^ > 0.7) or if no such variant was identified, the SNP was set to missing for that respective study. The study-specific ancestry-stratified summary statistics were first standardized to generate standard scores or normal deviates (z-scores), and then meta-analyzed using METAL in a fixed-effects model with inverse-variance weighting of regression coefficients^119^. In a first round of meta-analysis, ancestry-specific summary statistics were generated, which then served as input for a subsequent round of meta-analysis that represents the trans-ancestry effects of our lead SNPs on quantitative hepatic fat.

### External validation in a Liver Biopsy cohort

Available data from the following groups contributed to the Liver Biopsy Cohort.

#### Non-Alcoholic Steatohepatitis Clinical Research Network (NASH CRN) studies with Hispanic Boys, FLINT, PIVENS and NASH Women Studies

Results from several studies of EA and HISP participants were included from the Lundquist Institute. The Hispanic American cases are derived from the NAFLD Pediatric Database I (NAFLD Peds DB1), a prospective, longitudinal, multicenter, observational study cohort of adults and children initiated in 2002 and contains over 4,400 subjects^123^. Clinical and histologic features of database participants have been described by Patton et al^124^. Biopsy specimens were reviewed and scored centrally by the Non-Alcoholic Steatohepatitis Clinical Research Network (NASH CRN) Pathology Committee according to the histology scoring system established by the NASH CRN^125^. Genotyping was performed using the Illumina HumanCNV370-Quadv3 BeadChip at the Medical Genetics Institute at Cedars–Sinai Medical Center (HumanCNV370-Quadv3 BeadChips; Illumina, San Diego, CA, USA). The Hispanic American controls from NASH-CRN are derived from the Long QT Screening (LQTS) study^126^. Saliva samples were used for genotyping with the Illumina HumanCore-24 BeadChips at the Institute for Translational Genomics and Population Sciences of the Lundquist Institute at Harbor-UCLA Medical Center. For all Hispanic American samples, SNP data were imputed to the 1000 Genomes Project phase 3 dataset version 5 (AMR population) on the Michigan imputation server. The final dataset consisted of 787 samples, including 208 cases from NASH Boys and 579 controls from LQTS, and the top 3 PCs were included in the association analysis. The NASH CRN database and clinical trials were reviewed and approved by the individual institutional review boards at each participating site. All participants signed an informed consent prior to their enrollment into these consents and their de-identitied genetic data to be used for future liver disease research by the NASH CRN investigators and by their collaborators. These studies have been monitored by an NIDDK-sponsored data safety and monitoring board.

The European American NAFLD samples from Lundquist Institute are derived from FLINT, PIVENS, and NASH women studies, and the controls are derived from the Cholesterol and Atherosclerosis Pharmacogenetics (CAP) trial. The details of the <u>FLINT study</u> have been published previously^127^. Liver histology was blindly and centrally assessed by the NASH Clinical Research Network (NASH CRN) Pathology Committee according the NASH CRN system^125^. A total of 244 patients with available DNA were genotyped of whom 198 (81%) were White^128^. Genotyping was performed using the Omni2.5 content GWAS chip. <u>The PIVENS study</u>, a study of Pioglitazone versus Vitamin E versus Placebo in non-diabetic adults has been described previously^71^. Genotyping was performed on 432 PIVENS samples along with the FLINT samples using the Omni2.5 content GWAS chip. Subjects were removed for failed genotyping, unresolvable gender discrepancies, being outliers by principal component analyses, and by relatedness. In total, 197 White samples remained in the analysis dataset. <u>The NASH Women study</u> included a subset of patients who were into the NAFLD Database Study of NASH CRN whose liver biopsy specimens were reviewed and scored centrally by the NASH CRN Pathology Subcommittee. For the GWAS ancillary study^22^ genotyping was performed at the Medical Genetics Institute at Cedars–Sinai Medical Center with the use of Infinium HD technology (HumanCNV370-Quadv3 BeadChips; Illumina, San Diego, CA). The controls for the European American analysis from Lundquist Institute are derived from the CAP trial involved 944 healthy volunteers, 609 of whom were Caucasian^129^. In total, 591 subjects were genotyped on the Illumina HumanHap300 BeadChip or Illumina HumanCNV610-Quad beadchip. Imputation was performed using the Michigan Imputation Server with the reference panel of the Haplotype Reference Consortium (HRC) 1.1 release in 2016. After final QC of European American cohorts, the final 1,225 samples in the analysis including 650 cases and 575 controls, and the top 3 PCs for genetic ancestry were included in the association analysis.

#### EPoS Consortium Cohort

Results from EPoS consortium cohort were included from Newcastle University. A total 1,483 histologically characterized NAFLD cases were included and 17,781 genetically matched controls, with the cases recruited into the European NAFLD Registry (ClinicalTrials.gov Identifier: NCT04442334) from clinics at several leading European tertiary liver centres^20^. Details of inclusion/exclusion criteria have previously been described^20, 130^. All patients had undergone liver biopsy as part of the routine diagnostic workup for presumed NAFLD, and routinely assessed according to accepted criteria by experienced liver pathologists and scored using the well validated NIDDK NASH-CRN system^125^. Genotyping was performed using the Illumina OmniExpress BeadChip by Edinburgh Clinical Research Centre. The 17,781 population controls were recruited from existing genome-wide genotype data: Wellcome Trust Case Control Consortium, (n=5,159) typed on the Illumina Human1.2M-Duo; the Hypergenes cohort (n=1,520) typed on the Illumina Human1M-Duo, KORA (n=1,835) genotyped on the Illumina HumanOmni2.5 Exome chip, and Understanding Societies (n=9,267) typed on the Illumina HumanCoreExome chip. Overlapping SNPs that were well genotyped in all case and control cohorts were imputed together to the Haplotype Resource Consortium panel (HRC 1.1r 2016) by the Michigan Imputation Server.

#### The Geisinger Health System (GHS) bariatric surgery cohort

This consisted of 3,599 individuals of European American descent. Wedge biopsies of the liver were obtained intraoperatively during bariatric surgery, and liver histology was conducted by an experienced pathologist and subsequently re-reviewed by a second experienced pathologist using the NASH CRN scoring system^125^. A total of 806 participants did not have NAFLD and were classified as controls, whereas 2,793 were histologically characterized as having NAFLD. DNA sample preparation and whole-exome sequencing were performed at the Regeneron Genetics Centre^131^. Exome capture was performed using NimbleGen probes according to the manufacturer’s recommended protocol (Roche NimbleGen) multiplexed samples were sequenced on an Illumina v4 HiSeq 2500. Raw sequence data from each run were uploaded to the DNAnexus platform for sequence read alignment and variant identification.

#### STELLAR-3 and ATLAS studies

Results from two trials from Gilead Sciences were included including phase 3 STELLAR-3 study (ClinicalTrials.gov Identifier: NCT03053050), and phase 2 ATLAS study (ClinicalTrials.gov Identifier: NCT03449446) which were discontinued/terminated^132^. Genotyping was performed using whole genome sequencing (Illumina) aimed at 100x coverage. The PyVCF script was used to extract allele frequencies from VCF files generated using GATK4 pipeline with hg38 as reference genome.

#### BioVU Biorepository

BioVU subjects at Vanderbilt University underwent SNP genotyping using the Illumina Infinium Multi-Ethnic Genotyping Array (MEGAEX) platform and underwent QC analyses and imputation as previously described ^133^. Genetic data for were imputed using the Michigan Imputation Server (HRC v1.1) and genotyping data was linked to de-idenified EHR data. All available lab measurements in this cohort that occurred when the subject was at least 18 years of age. BioVU participants were selected based on available pathology report for liver biopsy in the note table in Observational Medical Outcomes Partnership (OMOP), excluding those with other conflicting diagnoses (e.g. viral hepatitis, alcohol, transplant, explant). NAFLD was defined based on pathology report defining hepatic fat as mild, moderate, severe, 5% or more.

Control subjects within BIoVU were identified by selecting those with ALT levels below 30 for males and below 20 for females. Both cases and controls were excluded for alcohol use disorders using ICD-9 and -10 codes.

#### Penn Medicine Biobank (PMBB)

The Penn Medicine Biobank includes participants recruited from the University of Pennsylvania Health System. A total of 139 biopsy proven NAFLD cases were selected using Linguamatics natural language processing on biopsy protocols of the PENN EHR. Cases were then linked to the PennMedicine BioBank. In addition, 1,995 PMBB participants were classified as controls if a recent CT scan of the liver was available, but no steatosis was present. Appropriate consent was obtained from each participant regarding storage of biological specimens, genetic sequencing, and access to all available EHR data. DNA extracted from the blood plasma of 2,134 samples were genotyped in three batches: the Illumina QuadOmni chip at the Regeneron Genetics Center; the Illumina GSA V1 chip OR on the Illumina GSA V2 chip by the Center for Applied Genomics at the Children’s Hospital of Philadelphia. Genotypes for each of the three PMBB datasets were imputed to the 1000 Genomes reference panel (1000G Phase3 v5) using the Michigan Imputation Server. Results from liver biopsy data are shown in **Supplemental Tables 6 and 7**.

### Heritability estimates and genetic correlations analysis

LD-score regression was used to estimate the heritability coefficient, and subsequently population and sample prevalence estimates were applied to estimate heritability on the liability scale^134^. A genome-wide genetic correlation analysis was performed to investigate possible co-regulation or a shared genetic basis between cALT and other complex traits and diseases (**Supplementary Table 9**). Pairwise genetic correlation coefficients were estimated between the meta-analyzed NAFLD GWAS summary output in EA and each of 774 precomputed and publicly available GWAS summary statistics for complex traits and diseases by using LD score regression through LD Hub v1.9.3 (http://ldsc.broadinstitute.org). Statistical significance was set to a Bonferroni-corrected level of *P* < 6.5 x 10^-5^.

### Tissue- and epigenetic-specific enrichment of NAFLD heritability

We analyzed cell type-specific annotations to identify enrichments of NAFLD heritability as shown in **Supplementary Table 16**. First, a baseline gene model was generated consisting of 53 functional categories, including UCSC gene models, ENCODE functional annotations^135^, Roadmap epigenomic annotations^136^, and FANTOM5 enhancers^137^. Gene expression and chromatin data were also analyzed to identify disease-relevant tissues, cell types, and tissue-specific epigenetic annotations. We used LDSC^37–39^ to test for enriched heritability in regions surrounding genes with the highest tissue-specific expression. Sources of data that were analyzed included 53 human tissue or cell type RNA-seq data from GTEx^28^; human, mouse, or rat tissue or cell type array data from the Franke lab^138^; 3 sets of mouse brain cell type array data from Cahoy *et al*^139^; 292 mouse immune cell type array data from ImmGen^140^; and 396 human epigenetic annotations from the Roadmap Epigenomics Consortium ^136^. Expression profiles are considered statistically significantly enriched for cALT susceptibility if they pass the nominal P-value threshold of 0.003.

### Pathway Annotation enrichment

Enrichment analyses in DEPICT^141^ were conducted using genome-wide significant (P < 5×10^-8^) NAFLD GWAS lead SNPs (**Supplementary Table 18**). DEPICT is based on predefined phenotypic gene sets from multiple databases and Affymetrix HGU133a2.0 expression microarray data from >37k subjects to build highly-expressed gene sets for Medical Subject Heading (MeSH) tissue and cell type annotations. Output includes a P-value for enrichment and a yes/no indicator of whether the FDR q-value is significant (P < 0.05). Tissue and gene-set enrichment features are considered. We tested for epigenomic enrichment of genetic variants using GREGOR software (**Supplementary Table 19**) ^142^. We selected EA-specific NAFLD lead variants with a p-value less than 5×10^−8^. We tested for enrichment of the resulting GWAS lead variants or their LD proxies (r^2^ threshold of 0.8 within 1 Mb of the GWAS lead, 1000 Genomes Phase I) in genomic features including ENCODE, Epigenome Roadmap, and manually curated data (**Supplemental Table 20**). Enrichment was considered significant if the enrichment p-value was less than the Bonferroni-corrected threshold of P=1.8×10^−5^ (0.05/2,725 tested features).

### Coding variant mapping

All imputed variants in MVP were evaluated with Ensemble variant effect predictor^143^, and all predicted LoF and missense variants were extracted. The LD was calculated with established variants for trans-ancestry, EA, AA, and HISP lead SNPs based on 1000 Genomes reference panel^144^. For SNPs with low allele frequencies, the MVP dataset was used for LD calculation for the respective underlying population. For the trans-ancestry coding variants, the EA panel was used for LD calculation. Coding variants that were in strong LD (r^2^ > 0.7) with lead SNPs and had a strong statistical association (P-value < 1×10^-5^) were considered the putative causal drivers of the observed association at the respective locus (**Supplementary Table 22**).

### Colocalization with gene expression

GWAS summary statistics were lifted over from GRCh37 to GRCh38 using LiftOver (https://genome.ucsc.edu/cgi-bin/hgLiftOver). Colocalization analysis was run separately for eQTLs and sQTLs for each of the 49 tissues in GTEx v8 (**Supplementary Tables 23 and 24**)^28^. For each tissue, we obtained an LD block for the genome with a sentinel SNP at P < 5×10^-8^, and then restricted analysis to the LD blocks. For each LD block with a sentinel SNP, all genes within 1Mb of the sentinel SNP (cis-Genes) were identified, and then restricted to those that were identified as eGenes in GTEx v8 at an FDR threshold of 0.05 (cis-eGenes). For each cis-eGene, we performed colocalization using all variants within 1Mb of the gene using the default prior probabilities in the ‘coloc’ function for the coloc package in R. We first assessed each coloc result for whether there was sufficient power to test for colocalization (PP3+PP4>0.8), and for the colocalization pairs that pass the power threshold, we defined the significant colocalization threshold as PP4/(PP3+PP4)>0.9.

### Overlap with open chromatin

At each of the 77 NAFLD-associated loci from the trans-ancestry meta-analysis, we looked for overlaps between any variant in the credible set, and regions of open chromatin previously identified using ATAC-Seq experiments in two cell types—3 biological replicates of HepG2^145^ and 3 biological replicates of hepatocyte-like cells (HLC)^146^ produced by differentiating three biological replicates of iPSCs, which in turn were generated from peripheral blood mononuclear cells using a previously published protocol^44^. Results are shown in **Supplementary Table 25**.

### Overlap with Promoter Capture-C data

We used two promoter Capture-C datasets from two cell/tissue types to capture physical interactions between gene promoters and their regulatory elements and genes; three biological replicates of HepG2 liver carcinoma cells, and hepatocyte-like cells (HLC)^145^. The detailed protocol to prepare HepG2 or HLC cells for the promoter Capture-C experiment is previously described^44^. Briefly, for each dataset, 10 million cells were used for promoter Capture-C library generation. Custom capture baits were designed using an Agilent SureSelect library design targeting both ends of DpnII restriction fragments encompassing promoters (including alternative promoters) of all human coding genes, noncoding RNA, antisense RNA, snRNA, miRNA, snoRNA, and lincRNA transcripts, totaling 36,691 RNA baited fragments. Each library was then sequenced on an Illumina NovoSeq (HLC), or Illumina HiSeq 4000 (HLC), generating 1.6 billion read pairs per sample (50 base pair read length.) HiCUP^147^ was used to process the raw FastQ files into loop calls; we then used CHiCAGO^148^ to define significant looping interactions; a default score of 5 was defined as significant. We identified those NAFLD loci at which at least one variant in the credible set interacted with an annotated bait in the Capture-C data (**Supplementary Table 25**).

### Protein-Protein Interaction Network Analysis

We employed the search tool for retrieval of interacting genes (STRING) v11^149^ (https://string-db.org) to seek potential interactions between nominated genes. STRING integrates both known and predicted PPIs and can be applied to predict functional interactions of proteins. In our study, the sources for interaction were restricted to the ‘Homo Sapiens’ species and limited to experimentally validated and curated databases. An interaction score > 0.4 were applied to construct the PPI networks, in which the nodes correspond to the proteins and the edges represent the interactions (**Figure 4, Supplemental Table 26**).

### Ensemble variant-to-gene mapping to identify putative causal genes

Based on DEPICT gene prediction, coding variant linkage analysis, QTL analysis, and annotation enrichment, and PPI networks **(Supplemental Tables 18-26**), a total of 215 potentially relevant genes for NAFLD were mapped to trans-ancestry 77 loci. For each locus, we counted how many times each gene in that region was identified in the 8 analyses. We then divided this number by the total number of experiments (i.e., 8) to calculate an evidence burden (called nomination score) that ranges from 0 to 100%. For each genomic locus, the gene that was most frequently identified as a causal gene was selected as the putative causal gene for that locus. In the case of a tie break, and if the respective genes have identical nomination profiles, the gene with eQTLs in multiple tissues was selected as the putative causal gene. Similarly, gene nomination was preferred for loci that strongly tagged (r^2^ > 0.8) a coding variant. Loci that scored with 3 distinct sources of evidence or greater are listed for coding variant (**Table 1A**) and non-coding variants (**Table 1B**), respectively.

### MVP LabWAS

A total of 21 continuous traits in the discovery MVP dataset, e.g. AST, ALP, fasting TG, HDL, LDL, TC, random glucose, HbA1c, albumin, bilirubin, platelet count, BMI, blood urea nitrogen (BUN), creatinine, eGFR, SBP, DBP, ESR, INR, and C-reactive protein were tested in 186,681 EA’s with association of 77 SNPs using linear regression of log-linear values. Covariates included age, gender and the first 10 PC’s of EA ancestry (**Supplementary Table 29**). The Bonferonni p-value threshold is set at 3.09×10^-05^ (0.05 / 21 traits * 77 SNPs)

### PheWAS with UK Biobank data

For the 77 lead trans-ancestry SNPs and EA and AA specific SNPs, we performed a PheWAS in a genome-wide association study of EHR-derived ICD billing codes from the White British participants of the UK Biobank using PheWeb^150^. In short, phenotypes were classified into 1,403 PheWAS codes excluding SNP-PheWAS code association pairs with case counts less than fifty^151^. All individuals were imputed using the Haplotype Reference Consortium panel^152^, resulting in the availability of 28 million genetic variants for a total of 408,961 subjects. Analyses on binary outcomes were conducted using a model named SAIGE, adjusted for genetic relatedness, gender, year of birth and the first 4 PC’s of white British genetic ancestry^153^. SAIGE stands for Scalable and Accurate Implementation of GEneralized mixed model and represents a generalized mixed-model association test that accounts for case-control imbalance and sample relatedness^153^. Results are shown in **Supplemental Tables 30 and 31**. SNP-trait associations are listed if they passed a nominal significance threshold of P < 0.001, and are considered Bonferoni significant when P < 4.6×10^-7^ (0.05 / 77 SNPs * 1,403 traits).

### IEU OpenGWAS project SNP lookup

An additional phenome-wide lookup was performed for 77 lead trans-ancestry SNPs and EA and AA specific SNPs in Bristol University’s MRC Integrative Epidemiology Unit (IEU) GWAS database^154^. This database consists of 126,114,500,026 genetic associations from 34,494 GWAS summary datasets, including UK Biobank (http://www.nealelab.is/uk-biobank), FinnGen (https://github.com/FINNGEN/pheweb), Biobank Japan (http://jenger.riken.jp/result), the NHGRI-EBI GWAS catalog (https://www.ebi.ac.uk/gwas), a large-scale blood metabolites GWAS^155^, circulating metabolites GWAS^156^, the MR-Base manually curated database^157^, and a protein level GWAS^158^. Results are shown in **Supplemental Table 32.**

### Regional cardiometabolic cross-trait colocalization

Bayesian colocalization tests between NAFLD-associated signals and the following trait- and disease-associated signals were performed using the COLOC R package^159^. To enable cross-trait associations, we compiled summary statistics of 36 cardiometabolic and blood cell-related quantitative traits and disease from GWAS studies conducted in EA ancestry individuals, and for MVP-based reports also on AA and HISP. To summarize, for total, HDL, and LDL cholesterol, triglycerides, alcohol use disorder, alcohol intake, systolic blood pressure, diastolic blood pressure, type 2 diabetes, BMI, CAD, we used the summary statistics available from various MVP-based studies^27, 116, 160^. Of these, the summary statistics for CAD and BMI GWAS in MVP have not been published or deposited as of yet. Data on WHR were derived from GIANT Consortium^161^, whereas summary statistics on CKD, gout, blood urea nitrogen, urate, urinary albumin-to-creatinine ratio, microalbuminuria, and eGFR were derived from CKD Genetics Consortium^162–164^. Finally, summary statistics of blood cell traits (e.g. platelet count, albumin, white blood cells, basophils, eosinophils, neutrophils, hemoglobin, hematocrit, immature reticulocyte fraction, lymphocytes, monocytes, reticulocytes, mean corpuscular hemoglobin, mean corpuscular volume, mean platelet volume, platelet distribution width, and red cell distribution width) were derived from a large-scale GWAS report performed in UK Biobank and INTERVAL studies^165^. A colocalization test was performed for all 77 NAFLD loci spanning 500kb region around the lead SNP for all 36 compiled traits. For each association pair COLOC was run with default parameters and priors. COLOC computed posterior probabilities for the following five hypotheses: PP0, no association with trait 1 (cALT GWAS signal) or trait 2 (e.g., co-associated metabolic signal); PP1, association with trait 1 only (i.e., no association with trait 2); PP2, association with trait 2 only (i.e., no association with trait 1); PP3, association with trait 1 and trait 2 by two independent signals; and PP4, association with trait 1 and trait 2 by shared variants. Evidence of colocalization^166^ was defined by PP3 + PP4 ≥ 0.99 and PP4/PP3 ≥ 5. Results are shown in **Supplemental Table 33.**

### Genetic risk scores and histologically characterized NAFLD

We constructed genetic risk scores (GRS) in 4 histologically characterized cohorts (e.g. Lundquist Whites and Hispanic Americans, EPoS Consortium Whites, and BioVU Whites) by calculating a linear combination of weights derived from the MVP dataset of lead 77 trans-ancestry cALT variants that passed conventional genome-wide significance (GRS-77, P < 5.0×10^-8^). The GRS-77 was standardized and the risk of histologically characterized NAFLD was assessed using a logistic regression model together with the potential confounding factors of age, gender, and the first 3 to 5 principal components of ancestry. To delineate the potential driving effects of known NAFLD loci, we divided the 77 loci into two sets, and generated one PRS consisting of 9 known NAFLD SNPs only (GRS-9), and one of newly identified 68 cALT SNPs (GRS-68). The goal of this separation is to evaluate whether a GRS based on novel SNPs alone (GRS-68) showed predictive capability for biopsy-proven histologically characterized NAFLD. Both GRS’s were added as independent predictors in a logistic regression model to explain histologically characterized NAFLD with the confounders of age, gender, and PC’s of ancestry. The individual effect sizes for each study were then meta-analyzed using the metagen package in R with random effects model comparing the standardized mean difference (SMD, mean differences divided by their respective standard deviations) **(Supplemental Table 9B)**. A forest plot was created to visualize the effect estimates between the studies **(Supplemental Figure 10)**. In similar fashion, SNPs were divided into 3 groups according to replication power, where SNPs were divided into a Bonferroni-replicated GRS consisting of 17 SNPs, a nominally significant with directional concordance GRS with 25 SNPs, and non-replicated GRS with 35 SNPs **(Supplemental Table 9C, Supplemental Figure 11).** Finally, a GRS subset was created based on the pleiotropy analysis and Venn Diagram, where we generated subset GRS that reflects liver+metabolic (17 SNPs), liver+metabolic+inflammation (38 SNPs), liver+inflammation (5 SNPs), and liver only strata (17 SNPs) **(Supplemental Table 9D, Supplemental Figure 14).**

### Transcription Factor Analysis

We identified nominated genes (**Supplemental Table 28**) that encode for TFs based on known motifs, inferred motifs from similar proteins, and likely sequence specific TFs according to literature or domain structure^167^. Target genes for these TFs were extracted using DoRothEA database^168^ in OmniPath collection^169^ using the associated Bioconductor R package OmnipathR^170^, a gene set resource containing TF-TF target interactions curated from public literature resources, such as ChIP-seq peaks, TF binding site motifs and interactions inferred directly from gene expression.

### Directional Pleiotropy and Gene Cluster Analysis

We used the R package ‘pheatmap’ for a stratified agglomerative hierarchical clustering method named ‘complete linkage’, where each element is its own cluster at the beginning, and two clusters of the shortest distance in between them are sequentially combined into larger clusters until all elements are included in one single cluster, where distance is measured in Euclidean distance. We used the 77 lead SNPs and their corresponding single-trait effect estimates for 20 traits corresponding to 3 biological super groups (e.g. lipids, inflammation, metabolic) as input, with the sign of each cell determined by direction of effect, and the strength by the –log10(p-value). The alleles were oriented as such that the cALT-increasing allele was set to the effect allele, which allows for direct comparison of the various association profiles. We selected the default ‘complete’ method and ‘Euclidean’ distance options to perform hierarchical clustering, stratified by the 3 super groups of metabolic, inflammation, and lipid traits. The results of the clustering gene set are visualized with a dendrogram on the left side of the heatmap, which is broadly grouped into 7 distinct gene clusters.

## Data Availability

The full summary level association data from the trans-ancestry, European, African American, Hispanic, and Asian meta-analysis from this report will be available through dbGAP project number phs001672.v4.p1 (study-specific accession codes will be available before publication).

https://www.ncbi.nlm.nih.gov/projects/gap/cgi-bin/analysis.cgi?study_id=phs001672.v4.p1

## Acknowledgements

This research is based on data from the Million Veteran Program, Office of Research and Development, Veterans Health Administration and was supported by award no. MVP000. This publication does not represent the views of the Department of Veterans Affairs, the US Food and Drug Administration, or the US Government. This research was also supported by funding from: the Department of Veterans Affairs awards I01- BX003362 (P.S.T. and K.M.C) and I01BX003341 (H.R.K. Co-Principal Investigator) and the VA Informatics and Computing Infrastructure (VINCI) VA HSR RES 130457 (S.L.D). B.F.V. acknowledges support for this work from the NIH/NIDDK (DK101478 and DK126194) and a Linda Pechenik Montague Investigator award. K.M.C, S.M.D, J.M.G, C.J.O, L.S.P, and P.S.T. are supported by the VA Cooperative Studies Program. S.M.D. is supported by the Veterans Administration [IK2 CX001780]. Funding support is also acknowledged for M.S. (K23 DK115897), R.M.C (R01 AA026302), D.K. (National Heart, Lung, and Blood Institute of the National Institutes of Health [T32 HL007734]), J.B.M (R01HL151855, UM1DK078616), L.S.P. (VA awards I01 CX001025, and I01 CX001737, NIH awards R21 DK099716, U01 DK091958, U01 DK098246, P30 DK111024, and R03 AI133172, and a Cystic Fibrosis Foundation award PHILLI12A0). The Rader lab was supported by NIH grants HL134853 (NJH and DJR) and DK114291-01A1 (K.T.C, N.J.H, and D.J.R). We thank all study participants for their contribution. Support for imaging studies was provided by ITMAT (NIH NCATS UL1TR001878), the Penn Center for Precision Medicine Accelerator Fund and R01 HL137501. Data for external replication and hepatic fat concordance were provided by investigators using United Kingdom BioBank, Multi-Ethnic Study of Atherosclerosis (MESA), Old Order Amish Study (Amish), Framingham Heart Study (FHS) and Penn Medicine Biobank (PMBB).

**MESA/MESA SHARe Acknowledgements**: MESA and the MESA SHARe projects are conducted and supported by the National Heart, Lung, and Blood Institute (NHLBI) in collaboration with MESA investigators. This research was supported by R01 HL071739 and MESA was supported by contracts 75N92020D00001, HHSN268201500003I, N01-HC-95159, 75N92020D00005, N01-HC-95160, 75N92020D00002, N01-HC-95161, 75N92020D00003, N01-HC-95162, 75N92020D00006, N01-HC-95163, 75N92020D00004, N01-HC-95164, 75N92020D00007, N01-HC-95165, N01-HC-95166, N01-HC-95167, N01-HC-95168, N01-HC-95169, UL1-TR-000040, UL1-TR-001079, UL1-TR-001420. Also supported in part by the National Center for Advancing Translational Sciences, CTSI grant UL1TR001881, and the National Institute of Diabetes and Digestive and Kidney Disease Diabetes Research Center (DRC) grant DK063491 to the Southern California Diabetes Endocrinology Research Center.

**NASH-CRN Acknowledgements:** The NASH Boys study was supported by NIDDK (U01DK061734, U01DK061718, U01DK061728, U01DK061731, U01DK061732, U01DK061737, U01DK061738, U01DK061730, U01DK061713) and NICHD. It was also supported by NIH CTSA awards (UL1TR000040, UL1RR024989, UL1RR025761, M01RR00188, UL1RR024131, UL1RR025014, UL1RR031990, UL1RR025741, UL1RR029887, UL1RR24156, UL1RR025055, UL1RR031980), and DRC HDK063491. LQTS was supported by the National Institutes of Health (grant 5R42HL112435-04 to QT Medical, Inc.). The provision of genotyping data was supported in part by the National Center for Advancing Translational Sciences, CTSI grant UL1TR001881, and the National Institute of Diabetes and Digestive and Kidney Disease Diabetes Research Center (DRC) grant DK063491 to the Southern California Diabetes Endocrinology Research Center. For LQTS: QT Medical, Inc., was involved in the study design and collection of data. However, QT Medical, Inc., had no involvement in the analysis and interpretation of the data, the drafting of the article, or the decision to submit the article for publication. The FLINT trial was conducted by the NASH CRN and supported in part by a Collaborative Research and Development Agreement (CRADA) between NIDDK and Intercept Pharmaceuticals. The Nonalcoholic Steatohepatitis Clinical Research Network (NASH CRN) is supported by the National Institute of Diabetes and Digestive and Kidney Diseases (NIDDK) (grants U01DK061718, U01DK061728, U01DK061731, U01DK061732, U01DK061734, U01DK061737, U01DK061738, U01DK061730, U01DK061713). Additional support is received from the National Center for Advancing Translational Sciences (NCATS) (grants UL1TR000439, UL1TR000436, UL1TR000006, UL1TR000448, UL1TR000100, UL1TR000004, UL1TR000423, UL1TR000058). The PIVENS study was supported by grants from the National Institutes of Health to the NASH Clinical Research Network (U01DK61718, U01DK61728, U01DK61731, U01DK61732, U01DK61734, U01DK61737, U01DK61738, U01DK61730, U01DK61713) and, in part, by the intramural program on the NIH, National Cancer Institute. Other grant support includes the following National Institutes of Health General Clinical Research Centers or Clinical and Translational Science Awards: UL1RR024989, UL1RR024128, M01RR000750, UL1RR024131, M01RR000827, UL1RR025014, M01RR000065. Additional funding to conduct PIVENS trial was provided by Takeda Pharmaceuticals North America, Inc. through a Cooperative Research and Development Agreement (CRADA) with the National Institutes of Health. The vitamin E softgels and matching placebo were provided by Pharmavite, LLC through a Clinical Trial Agreement with the National Institutes of Health. The NASH women study was supported by U01DK061737 and K24DK069290 (N.C.), NCRR grant M01-RR00425 to the Cedars-Sinai General Research Center Genotyping core, P30DK063491 to J.R., and R01DK079888 to M.O.G. This work is supported in part by the American Gastroenterological Association (AGA) Foundation, Sucampo, ASP Designated Research Award in Geriatric Gastroenterology, and by a T. Franklin Williams Scholarship Award; Funding provided by Atlantic Philanthropies, Inc, the John A. Hartford Foundation, the Association of Specialty Professors, and the American Gastroenterological Association to R.L. This research was funded in part with the support of the UCSD Digestive Diseases Research Development Center, US PHS grant DK080506.

The CAP study was supported by the National Institutes of Health: grant U19 HL069757 from the National Heart, Lung, and Blood Institute; and grant UL1TR000124 from the National Center for Advancing Translational Sciences.

**EPoS Acknowledgements:** The EPoS genetics study and the European NAFLD Registry have been supported by the EPoS (Elucidating Pathways of Steatohepatitis) consortium funded by the European Union Horizon 2020 Framework Program of the European Union under Grant Agreement 634413, the FLIP (Fatty Liver: Inhibition of Progression) consortium funded by the (European Union FP7 under grant agreement 241762), the LITMUS (Liver Investigation: Testing Marker Utility in Steatohepatitis) consortium funded by the European Union Innovative Medicines Initiative 2 Joint Undertaking that receives support from the European Union’s Horizon 2020 research and innovation programme and EFPIA under grant agreement 777377, and the Newcastle NIHR Biomedical Research Centre.

**Gilead Acknowledgements:** The Stellar and Atlas were funded by Gilead Sciences, Inc.

**Regeneron Acknowledgements:** The Geisinger Health System bariatric-surgery biobank was funded by Regeneron Pharmaceuticals and partly supported by a grant (P30DK072488) from the Mid-Atlantic Nutrition Obesity Research Center by the National Institutes of Health (NIH).

**Penn Medicine Biobank Acknowledgements:** The Penn Medicine BioBank is funded by the Perelman School of Medicine at the University of Pennsylvania and by a gift from the Smilow family, and the National Center for Advancing Translational Sciences of the National Institutes of Health under CTSA Award Number UL1TR001878.”

**BioVU Acknowledgements:** The Vanderbilt University Medical Center’s BioVU projects are supported by institutional funding, private agencies, and federal grants, which include the NIH funded Shared Instrumentation Grant S10OD017985 and S10RR025141; CTSA grants UL1TR002243, UL1TR000445, and UL1RR024975; and investigator-led projects U01HG004798, R01NS032830, RC2GM092618, P50GM115305, U01HG006378, U19HL065962, R01HD074711; U01 HG004603; U01 HG006378.

## Ethics statement

The Central Veterans Affairs Institutional Review Board (IRB) and site-specific Research and Development Committees approved the Million Veteran Program study. All other cohorts participating in this meta-analysis have ethical approval from their local institutions. All relevant ethical regulations were followed.

## Data availability

The full summary level association data from the trans-ancestry, European American, African American, Hispanic American, and Asian American meta-analysis from this report will be available through dbGAP (Accession codes will be available before publication).

