## Supplemental Figures for "A trans-ancestry genome-wide association study of unexplained chronic ALT elevation as a proxy for nonalcoholic fatty liver disease with histological and radiological validation"

### Slide 1
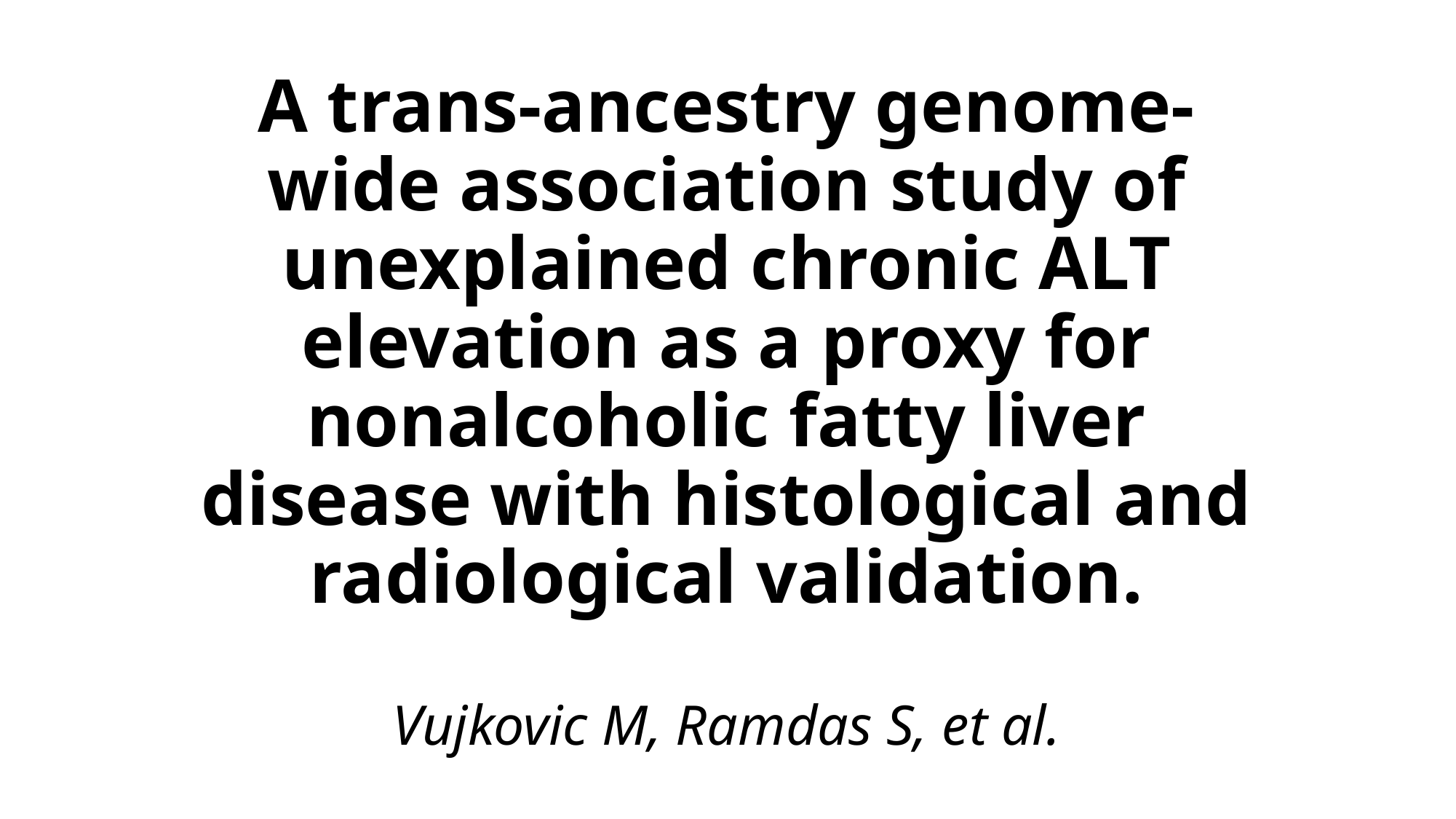

#### A trans-ancestry genome-wide association study of unexplained chronic ALT elevation as a proxy for nonalcoholic fatty liver disease with histological and radiological validation. Vujkovic M, Ramdas S, et al.

### Slide 2
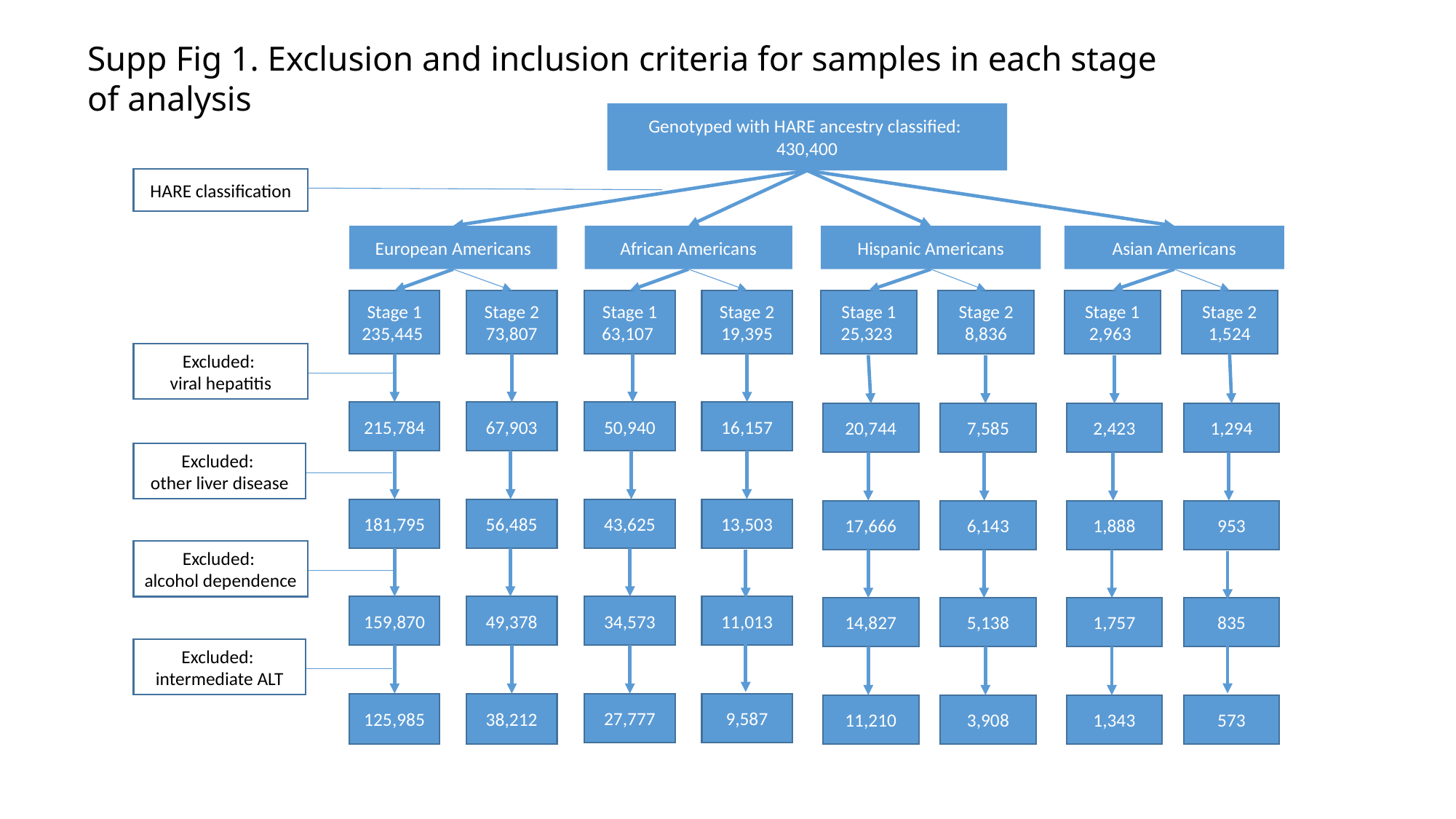

Supp Fig 1. Exclusion and inclusion criteria for samples in each stage of analysis
Genotyped with HARE ancestry classified:
430,400
HARE classification
European Americans
African Americans
Hispanic Americans
Asian Americans
Stage 1
235,445
Stage 2
73,807
Stage 1
63,107
Stage 2
19,395
Stage 1
25,323
Stage 2
8,836
Stage 1
2,963
Stage 2
1,524
Excluded:
viral hepatitis
215,784
67,903
50,940
16,157
20,744
7,585
2,423
1,294
Excluded:
other liver disease
181,795
56,485
43,625
13,503
17,666
6,143
1,888
953
Excluded:
alcohol dependence
159,870
49,378
34,573
11,013
14,827
5,138
1,757
835
Excluded:
intermediate ALT
125,985
38,212
27,777
9,587
11,210
3,908
1,343
573

### Slide 3
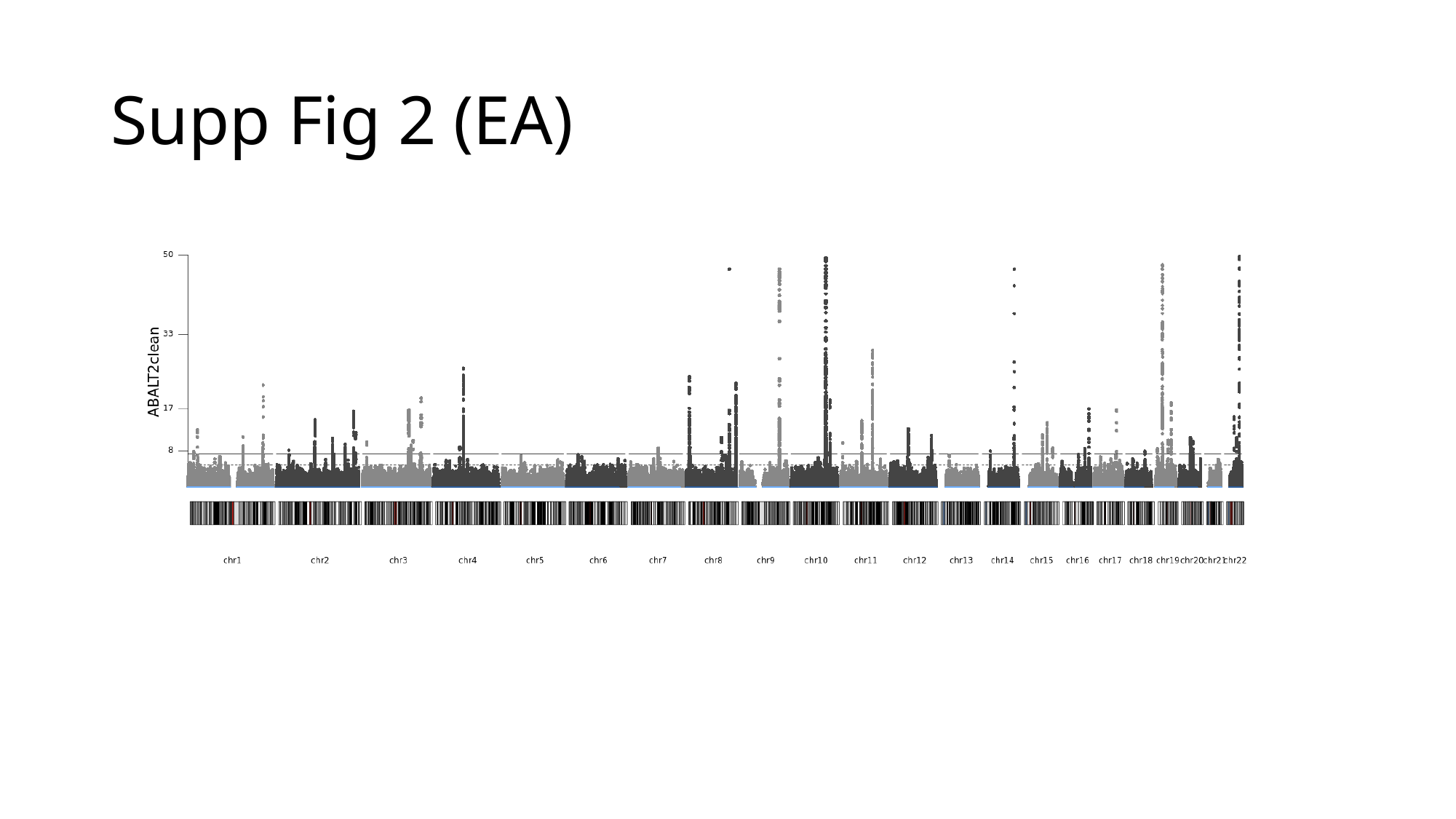

#### Supp Fig 2 (EA)

### Slide 4
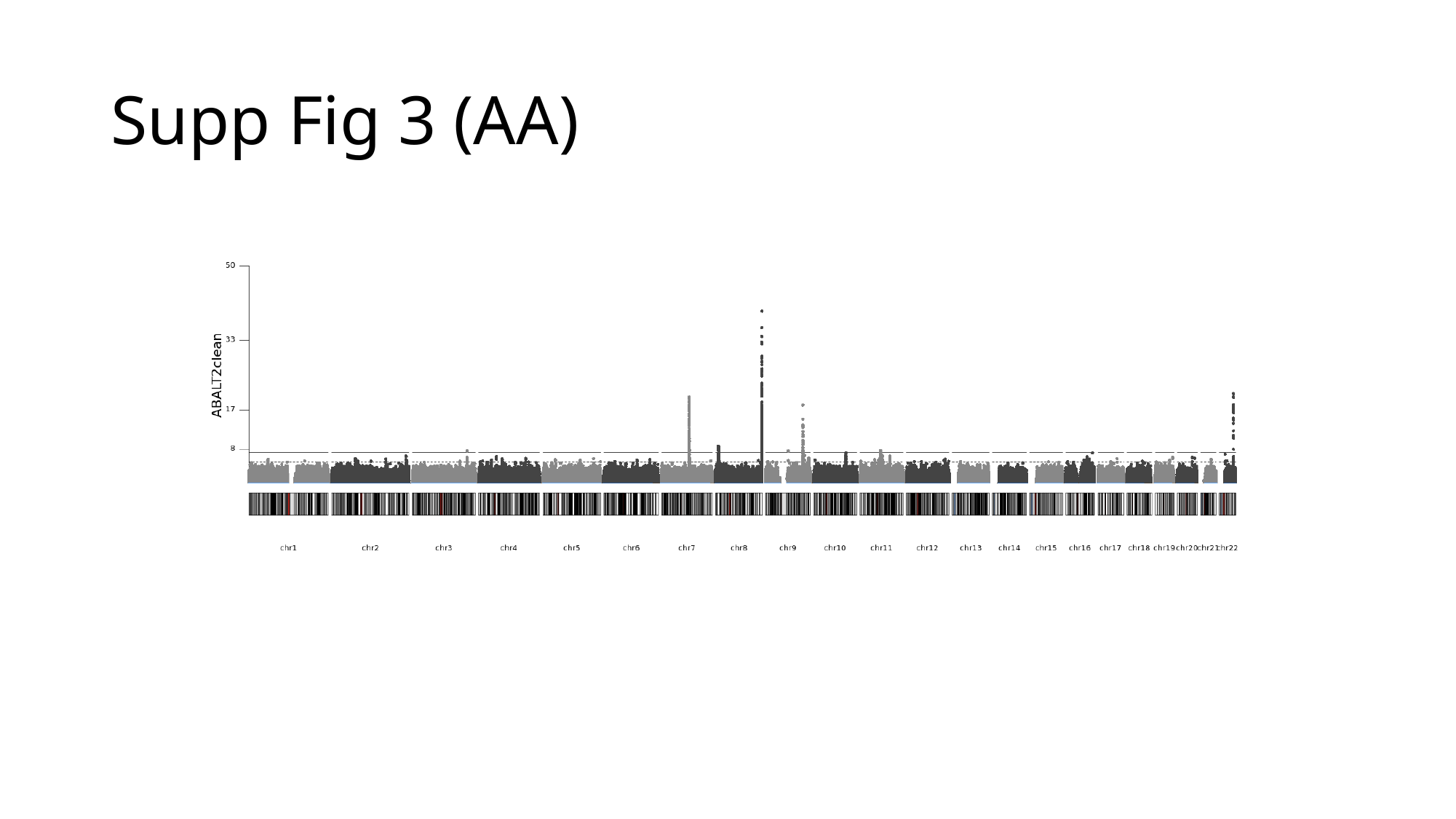

#### Supp Fig 3 (AA)

### Slide 5
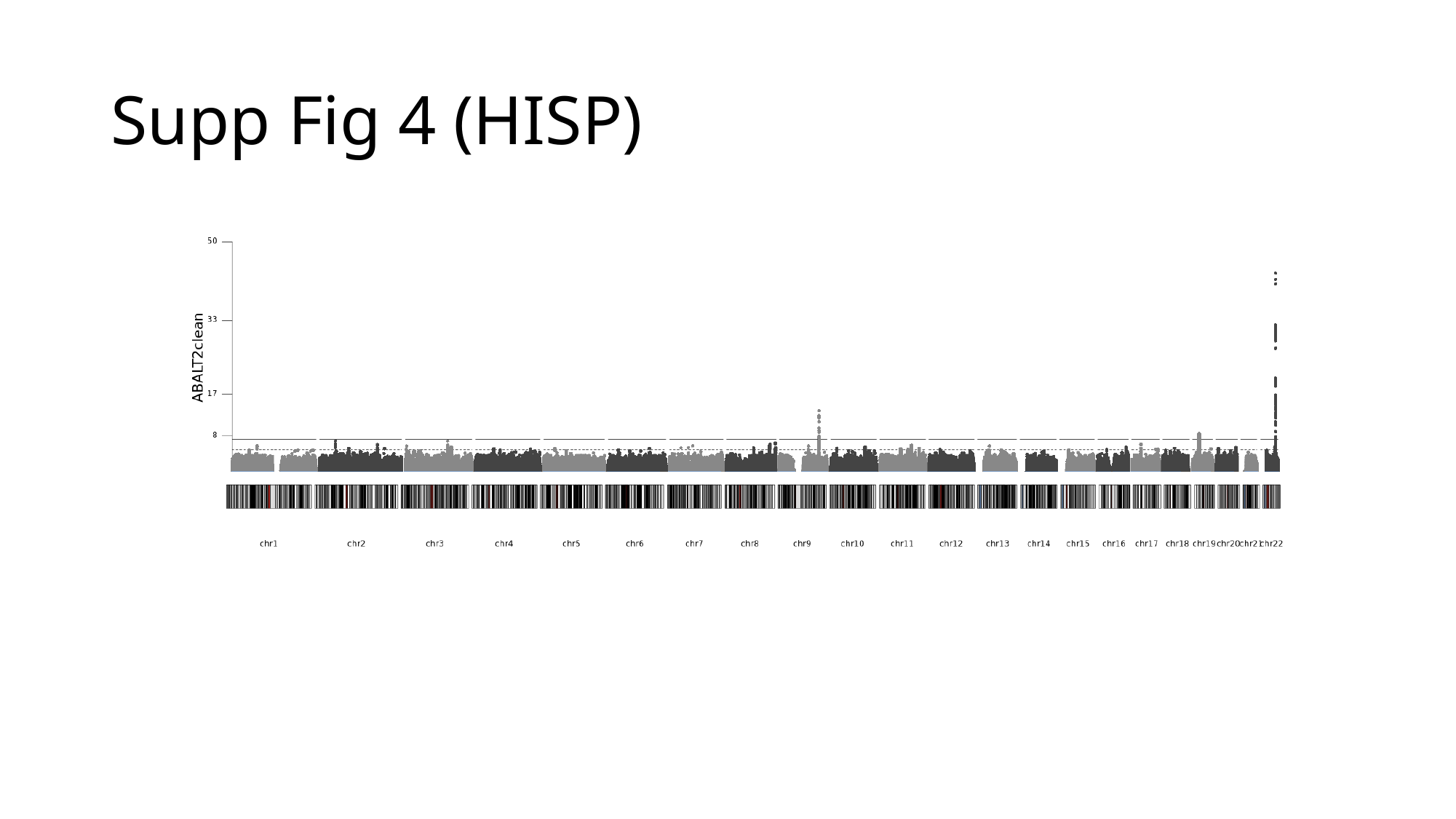

#### Supp Fig 4 (HISP)

### Slide 6
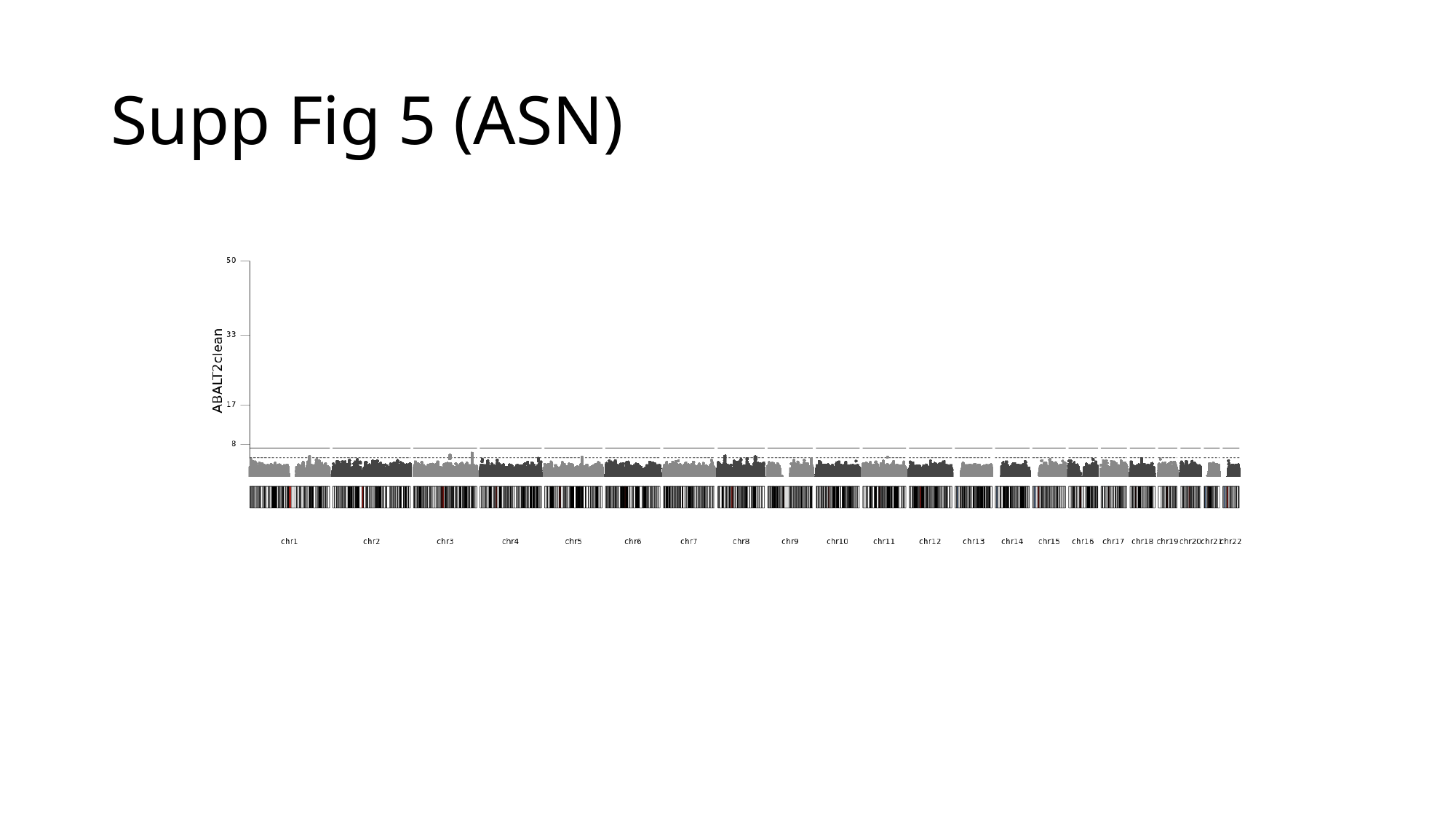

#### Supp Fig 5 (ASN)

### Slide 7
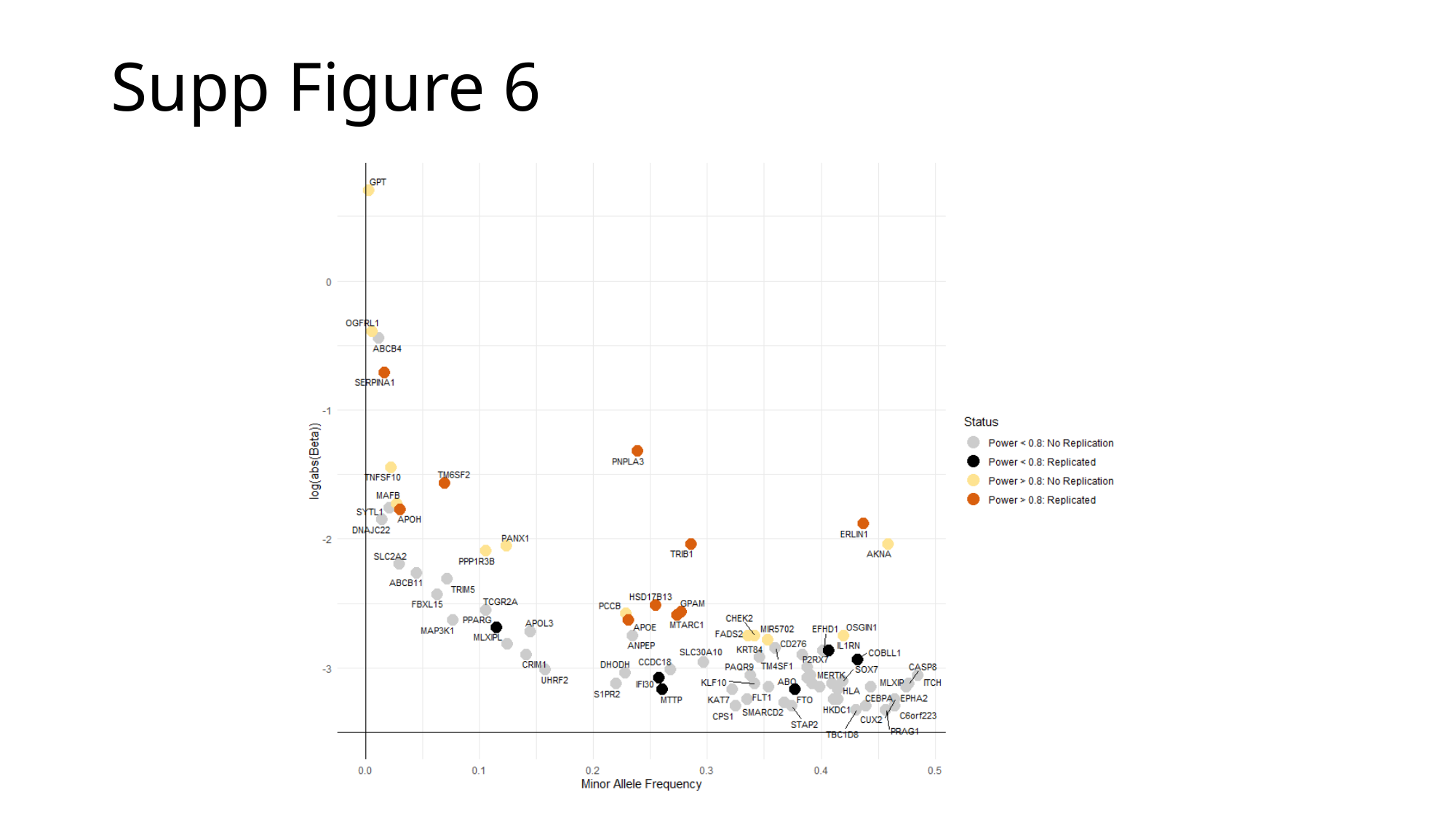

Supp Figure 6

### Slide 8
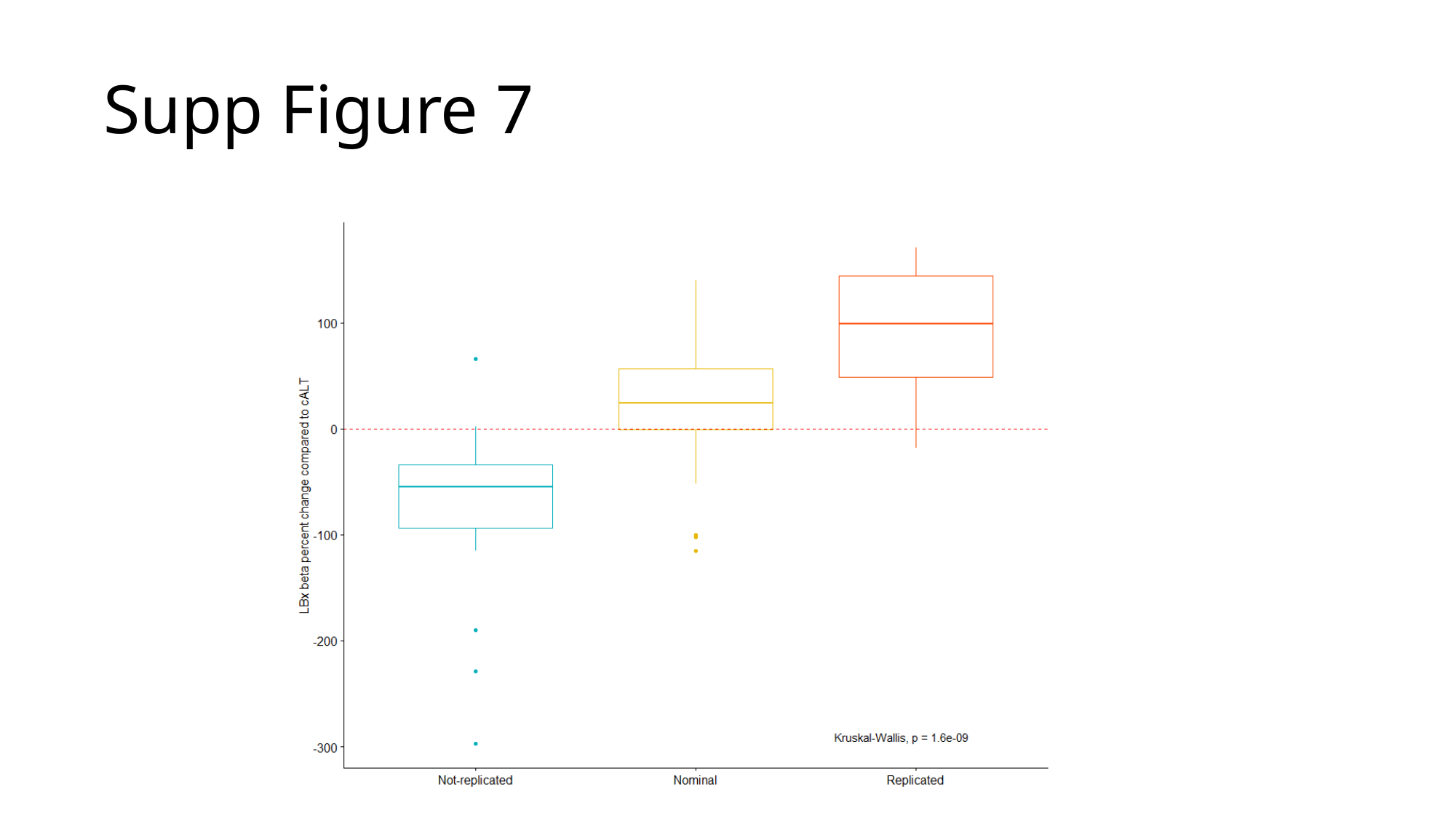

Supp Figure 7

### Slide 9
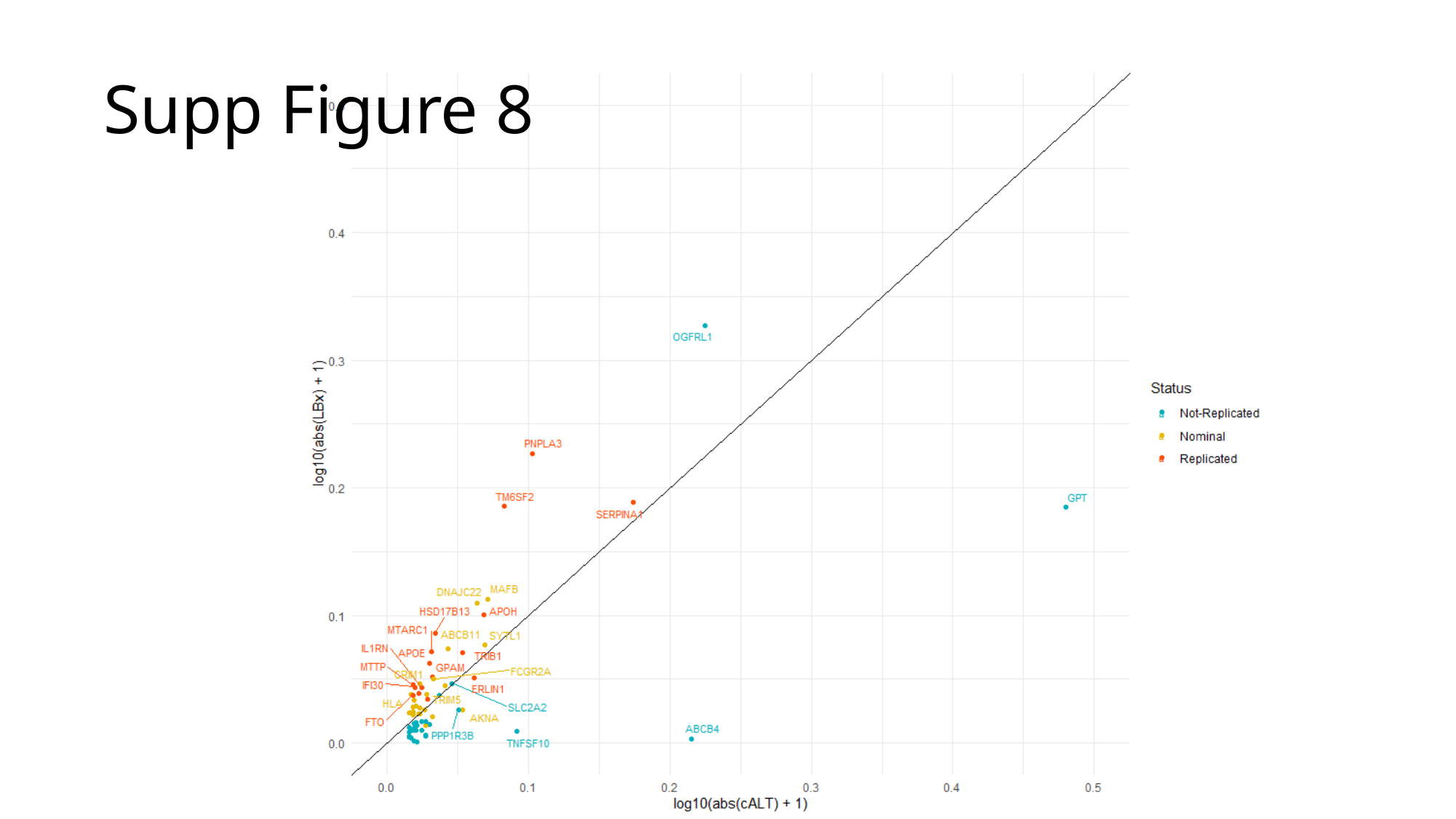

Supp Figure 8

### Slide 10
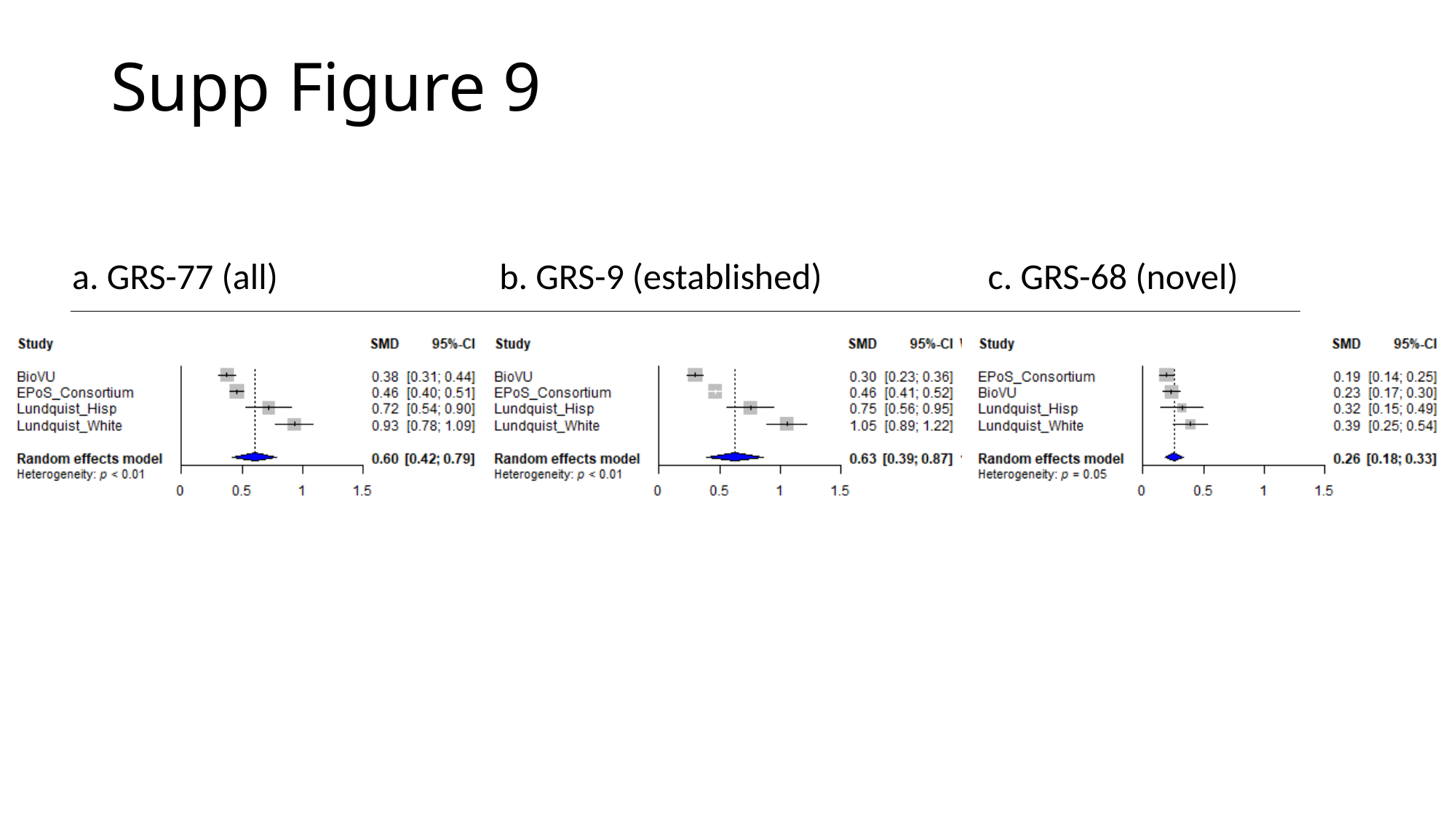

#### Supp Figure 9
b. GRS-9 (established)
c. GRS-68 (novel)
a. GRS-77 (all)

### Slide 11
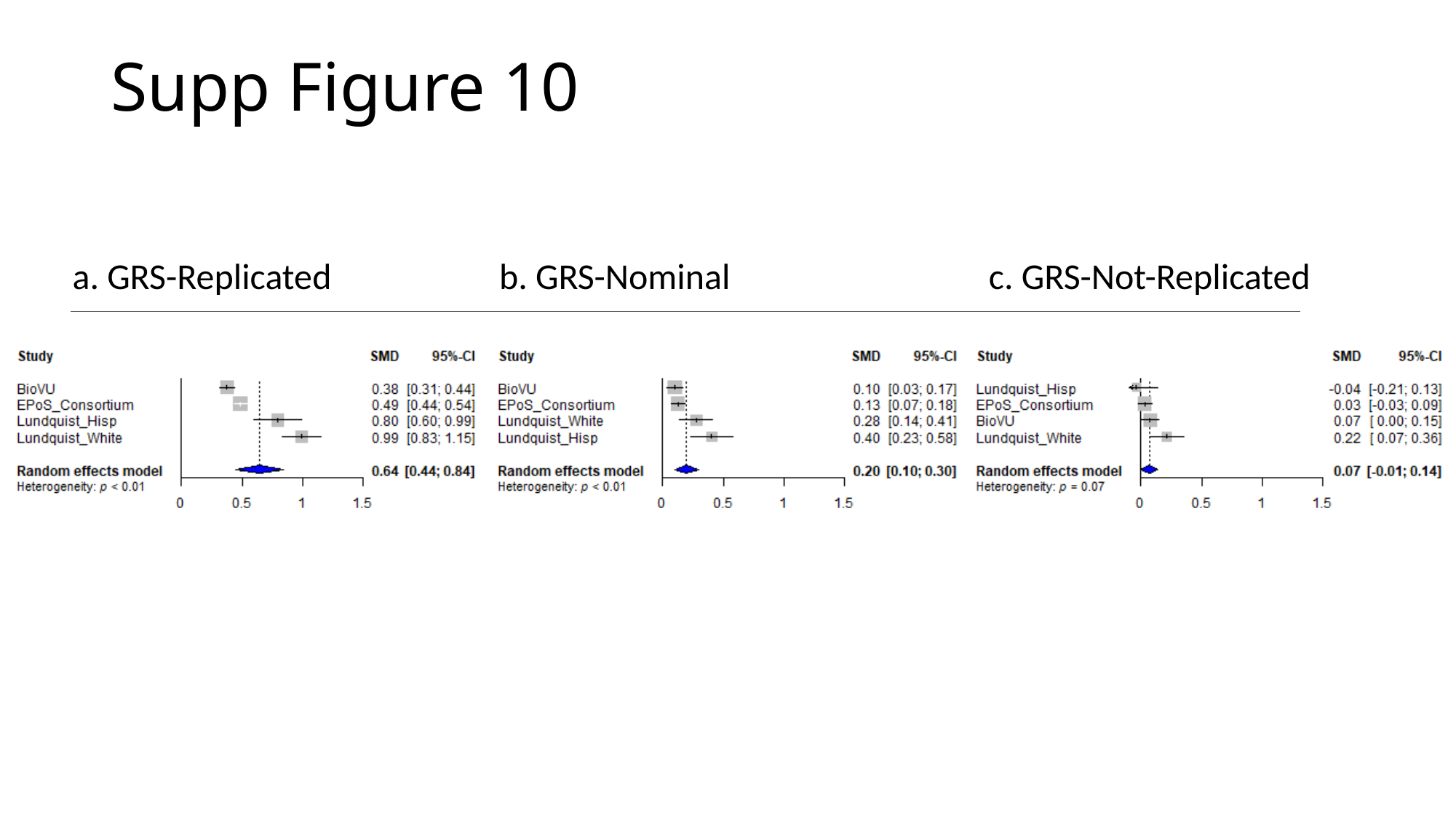

Supp Figure 10
b. GRS-Nominal
c. GRS-Not-Replicated
a. GRS-Replicated

### Slide 12
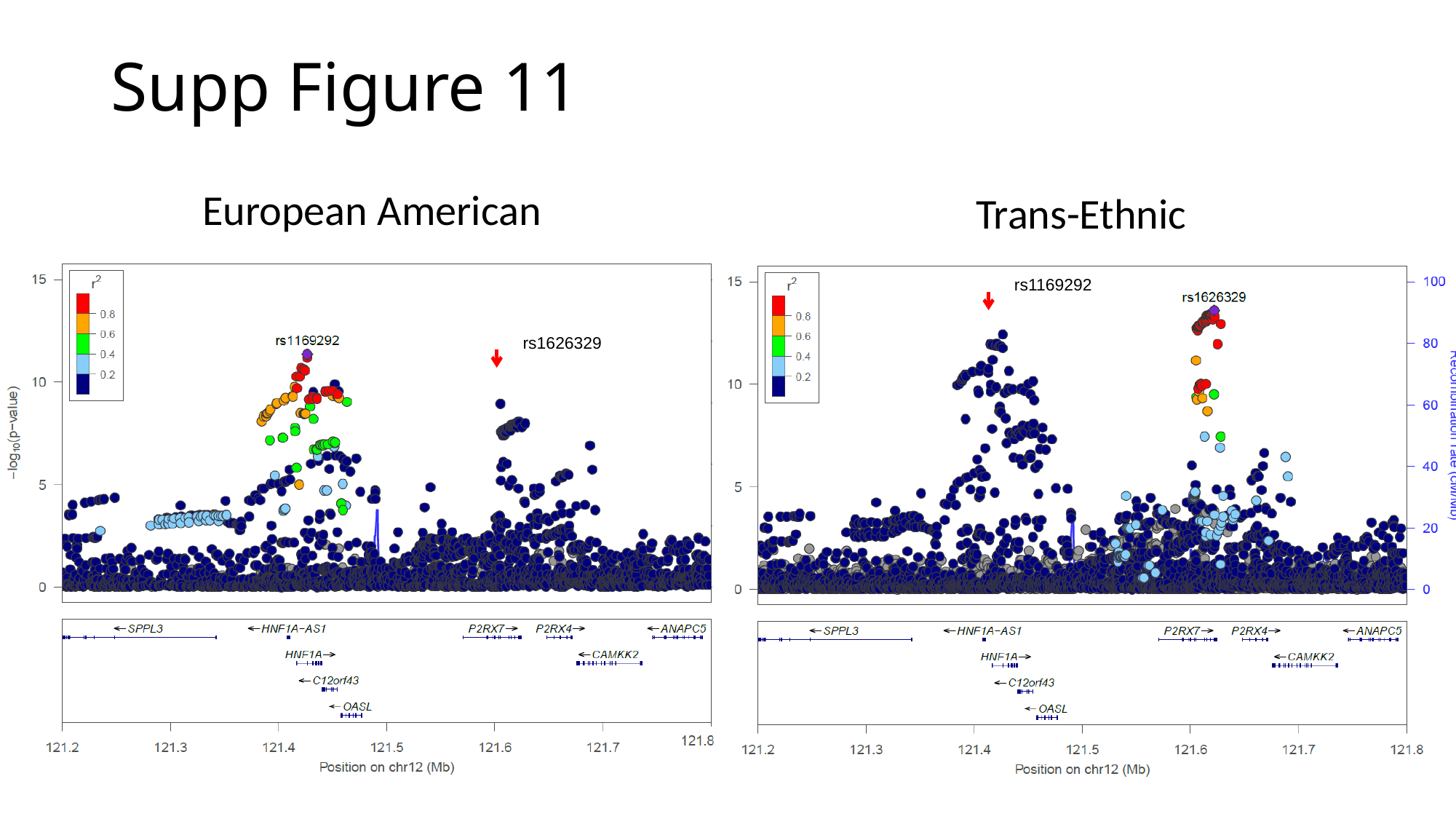

Supp Figure 11
European American
Trans-Ethnic
rs1169292
rs1626329

### Slide 13
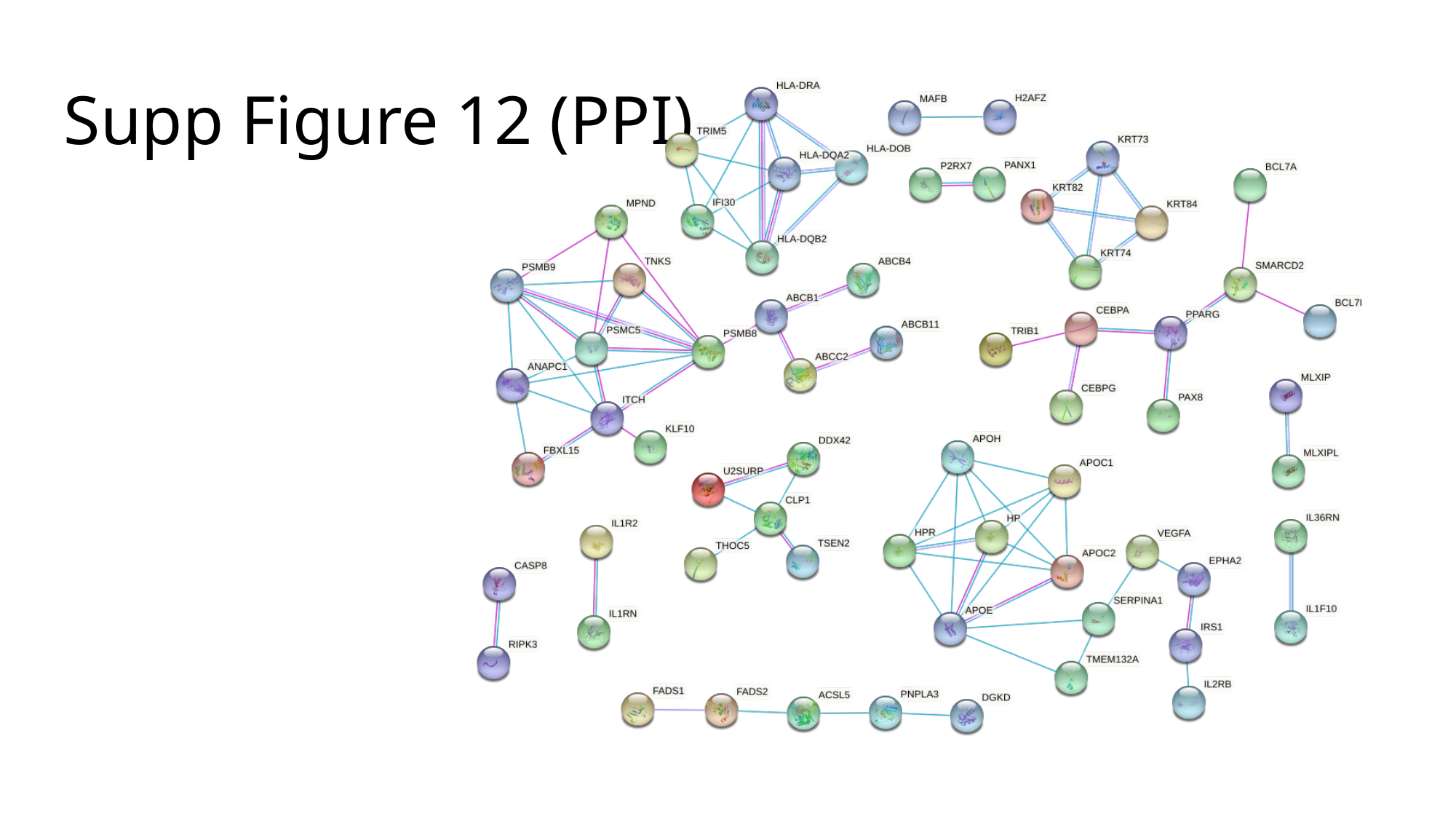

#### Supp Figure 12 (PPI)

### Slide 14
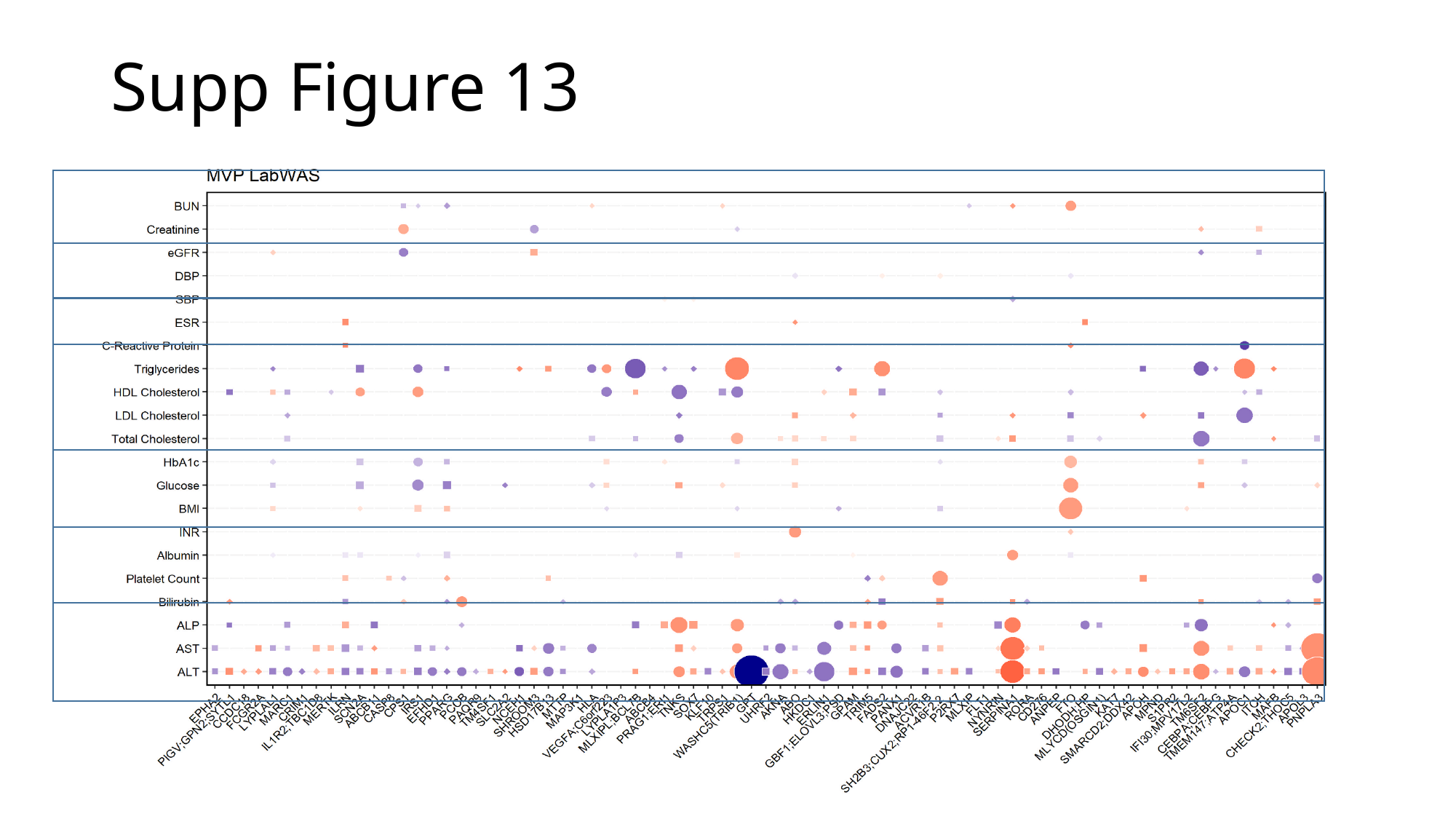

#### Supp Figure 13

### Slide 15
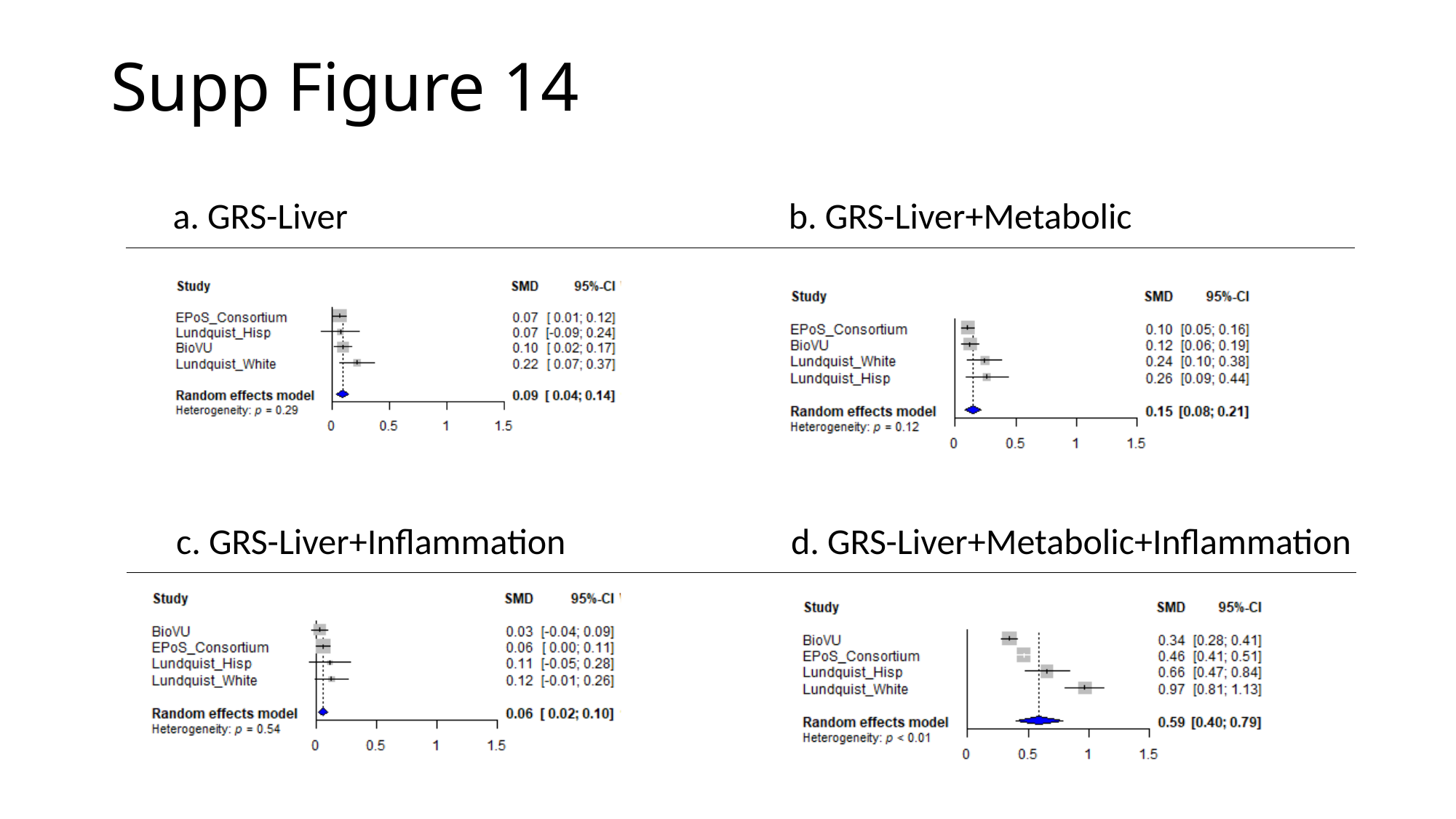

Supp Figure 14
b. GRS-Liver+Metabolic
a. GRS-Liver
d. GRS-Liver+Metabolic+Inflammation
c. GRS-Liver+Inflammation
